# Angiography-Derived Autoregulation Targets After Thrombectomy

**DOI:** 10.64898/2026.08.13.26360389

**Authors:** Kyle A. Lyman, Lily Pwint Thinzar, David Vargas, Guido J. Falcone, Emily J. Gilmore, Jennifer Ahjin Kim, Jessica R. Magid-Bernstein, Adam de Havenon, Charles Matouk, Ryan Hebert, Kevin N. Sheth, Santiago Ortega-Gutierrez, Nils H. Petersen

## Abstract

Optimal blood pressure management after thrombectomy remains uncertain, and individualized autoregulation-based targets typically require continuous neuromonitoring. We developed an angiography-derived autoregulatory metric using intraprocedural data and applied it retrospectively to a single-center cohort of patients who underwent thrombectomy for acute stroke. From 62 patients with 3-month functional outcomes, greater time within the predicted autoregulatory range during the first 24 hours after thrombectomy was independently associated with improved outcome after adjustment for covariates (odds ratio per 10% increase, 1.86; 95% CI, 1.31-2.66; P = .0006). These findings support routine angiography as a potential source of early, patient-specific hemodynamic targets after thrombectomy.

## INTRODUCTION

Optimal blood pressure management after endovascular thrombectomy remains uncertain.^1^ Both hypotension and hypertension may worsen secondary brain injury following reperfusion, yet current guidelines rely on population-based blood pressure thresholds rather than patient-specific cerebrovascular physiology.^2,3^ Emerging evidence suggests that individualized blood pressure targets based on cerebral autoregulation may improve outcomes after stroke.^4^ Near-infrared spectroscopy (NIRS) monitoring can estimate autoregulatory limits and optimal mean arterial pressure (MAP_Opt_), but the need for specialized equipment and prolonged monitoring limits scalability and may delay availability of actionable physiologic information during the early post-reperfusion period.^4,5^ We therefore asked whether routine angiography and intraprocedural physiologic data could be repurposed to generate an early, patient-specific estimate of autoregulatory target after thrombectomy. In this study, we developed an angiography-derived estimate of autoregulatory target and then tested whether postprocedural blood pressure exposure relative to that estimate was associated with 3-month functional outcome.

## SUBJECTS/MATERIALS AND METHODS

We conducted a retrospective secondary analysis of patients previously enrolled in a prospective observational study of anterior circulation large-vessel occlusion (LVO) treated with endovascular thrombectomy.^4^ The institutional review board approved the study and granted a waiver of informed consent. Reporting of this study conforms to the STROBE statement (checklist included as Supplementary). The analytic framework is summarized in Fig S1. We hypothesized that angiography-derived perfusion metrics would estimate a patient’s autoregulatory status because the relationships among MAP, tissue perfusion, and end-tidal CO_2_ (EtCO_2_) reflect core cerebrovascular physiology rather than a disease-specific feature of aneurysmal subarachnoid hemorrhage (aSAH) or LVO. We derived the mapping between angiographic perfusion and NIRS-defined autoregulatory state in an independent aSAH cohort with concurrent angiography and NIRS monitoring. The model estimated Scale_D_, a normalized measure of a patient’s MAP relative to their autoregulation-based MAP_Opt_ (Fig 1A). Details of the angiographic perfusion workflow are provided in Fig S2.^6,7^ Briefly, the post-thrombectomy lateral ICA angiographic acquisition from the reperfused hemisphere was temporally matched to the anesthesia record using DICOM timestamps, and MAP_Angio_ and EtCO_2_ were calculated by averaging the prespecified 5-minute interval preceding and including the acquisition. Second, angiographic signal-time curves were converted into pixel-wise perfusion estimates after exclusion of large vascular pixels and gamma-variate extrapolation when needed to calculate mean transit time (MTT) from the hemispheric tissue mask. Third, the aSAH derivation model used MTT, MAP_Angio_, EtCO_2_, age, sex, and interaction terms to estimate Scale_D_, which was then calibrated in the LVO cohort to predict MAP_Opt_. Only the reperfused hemisphere was considered for analysis because the contralateral hemisphere was not systematically examined as part of routine post-thrombectomy care. Cohort characteristics are summarized in Tables S1-2.

**Figure 1.**
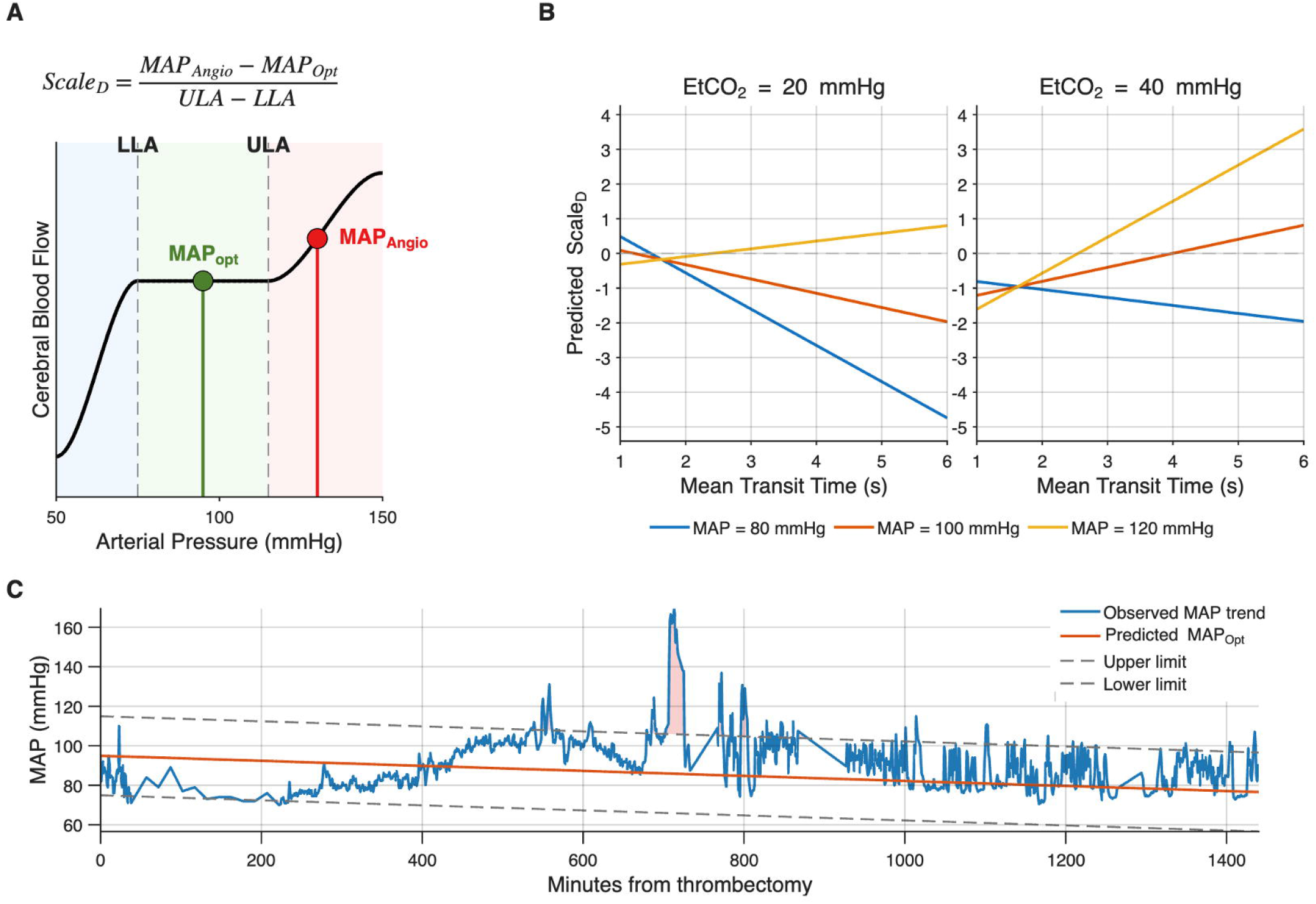
Estimation of Cerebral Autoregulatory Position from Angiography-Derived Physiologic Data. **A**, Conceptual schematic of the scaled autoregulatory-distance metric, *Scale*_*D*_, defined as the distance between mean arterial pressure during angiography (MAP_Angio_) and optimal mean arterial pressure (MAP_Opt_), normalized by the width of the autoregulatory range bounded by the lower and upper limits of autoregulation (LLA and ULA). Positive values indicate pressure above MAP_Opt_; negative values indicate pressure below MAP_Opt_. **B**, Model-derived relationships between angiography-derived mean transit time, MAP_Angio_, and estimated autoregulatory position at fixed end-tidal carbon dioxide values, illustrating the model-derived dependence of *Scale*_*D*_ on contemporaneous perfusion and physiologic conditions. **C**, Representative patient-level reconstruction of post thrombectomy blood pressure exposure over 24 hours. The observed minute-by-minute MAP trajectory is shown relative to the predicted MAP_Opt_ and the corresponding predicted autoregulatory limits. Shaded regions indicate excursions outside the predicted autoregulatory range, which were used to calculate the proportion of time spent within predicted limits. **Abbreviations:** EtCO_2_, end-tidal carbon dioxide; LLA, lower limit of autoregulation; MAP, mean arterial pressure; MAP_Opt_, optimal mean arterial pressure; MTT, mean transit time; ULA, upper limit of autoregulation.

A mixed-effects model incorporating demographic characteristics, angiographic perfusion, and intraprocedural physiologic variables was used to predict Scale_D_ (Fig 1B; Table S3; Figs S3-6). The model was calibrated in a subset of thrombectomy cases with high-quality angiography to estimate MAP_Opt_ from Scale_D_ and time from reperfusion (Table S4 and Figs S7-11). ^6,7^ For each patient, minute-by-minute MAP trajectories were reconstructed for the first 24 hours after reperfusion. The upper and lower autoregulatory limits were operationalized as predicted MAP_Opt_ ± 20 mmHg and the proportion of time spent within their predicted autoregulatory range was calculated (Fig 1C). The primary outcome was 3-month modified Rankin Scale (mRS) score. Associations between the proportion of time within the predicted autoregulatory range and outcome were evaluated using covariate-adjusted ordinal logistic regression. Sensitivity analyses included partial proportional-odds and dichotomous good-outcome models. Reperfusion status was included as an outcome-model covariate, but the Scale_D_-to-MAP_Opt_ calibration model was intentionally restricted to physiologic and angiographic inputs because the goal was to estimate autoregulatory state.

## RESULTS

Of 64 patients with analyzable angiographic data, 62 patients had 3-month outcome data and were included in the primary analysis. The distribution of 3-month mRS scores is shown in Table S6; 32 patients (51.6%) achieved functional independence (mRS 0-2). Descriptive characteristics stratified by time spent within the predicted autoregulatory range for the full cohort are provided in Table S5.

Greater time within the predicted autoregulatory range was associated with a favorable shift in 3-month mRS (Fig 2A). In the unadjusted analysis, this association was directionally favorable but not significant (Wald χ^2^ = 1.38, p = 0.24). In multivariable proportional-odds regression, greater time within the predicted autoregulatory range was independently associated with a favorable shift in 3-month mRS (odds ratio per 10% increase, 1.86; 95% CI, 1.31-2.66; P = .0006) (Fig 2B; Table S7). Older age, higher admission stroke severity, and greater baseline disability were associated with worse outcomes, whereas intravenous thrombolysis and better reperfusion were associated with improved outcomes. The model demonstrated good discrimination (c statistic, 0.81).

**Figure 2.**
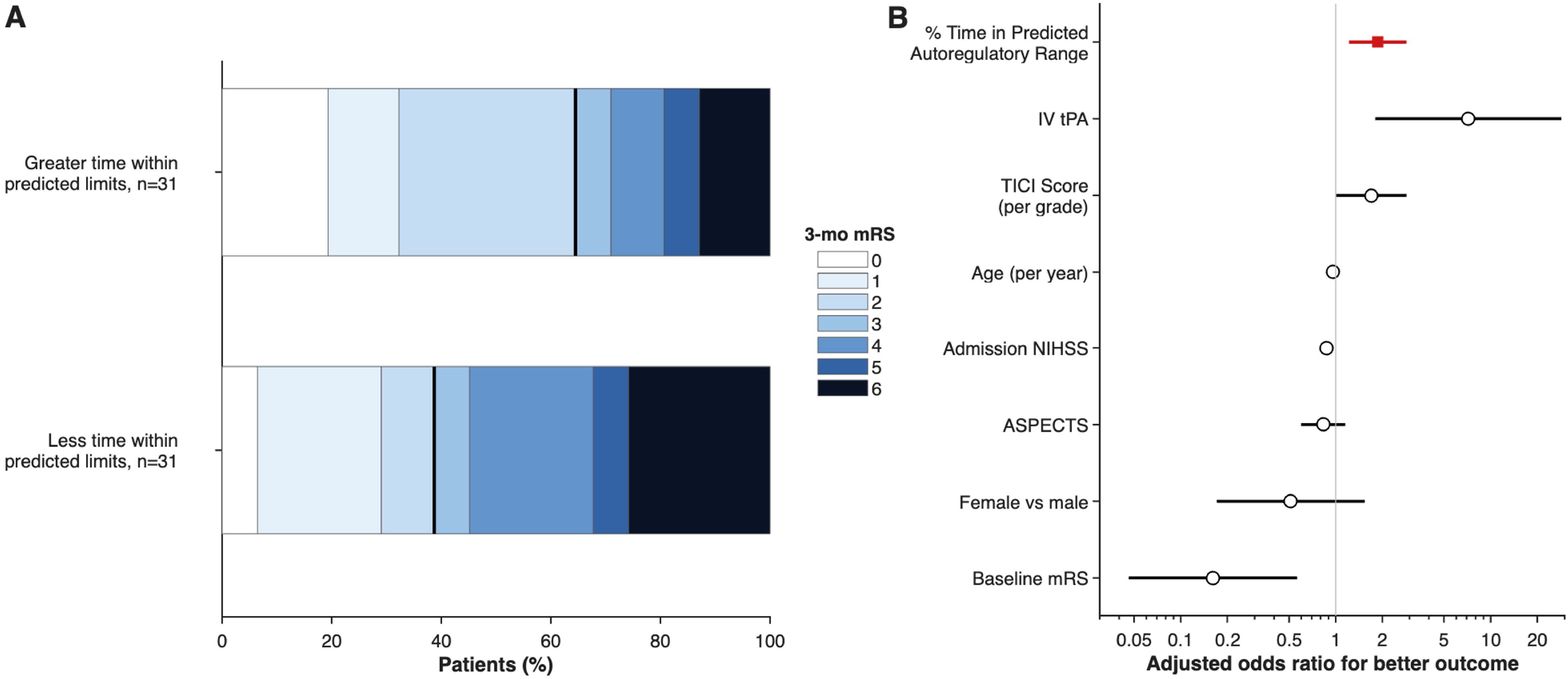
Predicted Autoregulatory Exposure and Functional Outcome After Thrombectomy. **A**, Distribution of 3-month modified Rankin Scale (mRS) scores in patients above vs below the cohort median proportion of time spent within the predicted autoregulatory range during the first 24 hours after thrombectomy. Patients with greater time within predicted limits showed a favorable shift in functional outcome distribution. Vertical lines denote the boundary between mRS 0 to 2 and mRS 3 to 6. **B**, Forest plot of adjusted odds ratios from the proportional-odds ordinal logistic regression model for 3-month mRS. Odds ratios are displayed so that values greater than 1 indicate a favorable shift toward lower 3-month mRS. The primary exposure, percentage of time in the predicted autoregulatory range, is shown in red. Estimates were adjusted for age, intravenous thrombolysis, TICI score, baseline mRS, admission NIHSS, ASPECTS, and sex. **Abbreviations:** ASPECTS, Alberta Stroke Program Early CT Score; mRS, modified Rankin Scale; NIHSS, National Institutes of Health Stroke Scale; TICI, Thrombolysis in Cerebral Infarction.

Sensitivity analyses were concordant with the primary findings and robust to changes in modeling assumptions (Fig S12). In a partial proportional-odds model, greater time within the predicted autoregulatory range remained significantly associated with mRS overall (Table S8). In a dichotomous model, greater time within the predicted autoregulatory range was independently associated with functional independence (odds ratio per 10% increase, 3.92; 95% CI, 1.44-10.68; P = .0075) (Table S9). When the physiology-derived Scale_D_ term was omitted, the association between autoregulatory exposure and functional outcome was no longer significant (Fig S13).

## DISCUSSION

In this proof-of-concept clinical application, routine angiography and intraprocedural physiologic data were used to generate an early, patient-specific estimate of post-thrombectomy autoregulatory target, and blood pressure exposure relative to that estimate was independently associated with 3-month functional outcome. The principal conceptual advance is not that angiography alone serves as a definitive bedside surrogate for continuous autoregulation monitoring, but that routinely acquired procedural data may contain enough physiologic signal to support early hemodynamic stratification after reperfusion. This framing is important because the method is most appropriately interpreted as a physiology-informed estimator with clinical relevance, rather than as a point-precise replacement for NIRS-derived MAP_Opt_.

Prior autoregulation-guided approaches have depended on continuous NIRS monitoring, which may be unavailable in many centers and often becomes informative only after a substantial delay.^4,5^ By contrast, the present framework leverages data already acquired during thrombectomy to estimate an autoregulatory target immediately after reperfusion, thereby extending physiologic assessment into an early window that may be particularly vulnerable to secondary injury. The observed association between greater time within the predicted autoregulatory range and better outcome supports the clinical relevance of that early estimate, while the model’s moderate patient-level precision argues for prospective refinement and validation before it is used to direct care.

The observed association between autoregulatory exposure and outcome is consistent with the hypothesis that individualized blood pressure targets may improve neurological recovery following reperfusion therapy.^8,9^ Patients whose blood pressure remained within their predicted autoregulatory limits had better outcomes even after adjustment for established predictors of stroke recovery. At the same time, residual confounding remains possible, because time within the predicted autoregulatory range may also partly reflect other favorable but incompletely measured features of recovery, including greater hemodynamic stability, less severe reperfusion injury, smaller infarct burden, or more consistent post-thrombectomy critical care. This approach may also help to reconcile conflicting evidence in post-thrombectomy blood pressure management by shifting from uniform thresholds to patient-specific targets aligned with underlying cerebrovascular physiology.^10,11^ Beyond clinical associations, these findings may support the feasibility of inferring autoregulatory state from routine procedural data. By linking angiographic perfusion metrics with intraprocedural physiologic signals, this approach may extend autoregulation-guided care from a monitoring-dependent paradigm to one that may be more broadly implementable.

This study has several limitations. First, it was retrospective and single-center.^4^ Second, the physiologic model was derived in an independent cohort of patients with aSAH and, although calibrated in ischemic stroke, requires prospective validation. Third, estimation of autoregulatory limits relied on simplified modeling assumptions that may not capture dynamic cerebrovascular physiology. Time within the predicted autoregulatory range may reflect physiologic stability or quality of care rather than a mechanistic treatment target. Accordingly, these findings support a clinically meaningful association but do not show that maintaining blood pressure within the predicted range improves recovery. The modest sample size limited the ability to perform well-powered subgroup analyses by occlusion location, degree of reperfusion, or contralateral versus ipsilateral physiology. The angiographic MTT workflow has been evaluated in prior work for feasibility and truncation behavior in DSA^6,7^, but it was not validated against contemporaneous CT or MR perfusion in the present LVO cohort. The derivation cohort also consisted of patients with aSAH, who differ from patients with ischemic LVO in age, vascular injury mechanism, and temporal evolution of autoregulation; this disease-state transportability is therefore an important limitation and a target for prospective validation. Finally, the model was derived from NIRS-based MAP_Opt_ estimates, and NIRS itself has important limitations, including limited spatial sampling and susceptibility to artifacts.

Routine angiography may provide an early, physiology-informed estimate of patient-specific autoregulatory target after thrombectomy, and blood pressure exposure relative to that estimate was associated with functional outcome in this retrospective cohort. These findings support prospective real-time validation and interventional testing of angiography-guided blood pressure management after reperfusion.

## Data Availability

All data produced in the present study are available upon reasonable request to the authors

## ACKNOWLEDGEMENTS

This work was supported by the NIH and the AHA, but the contents are solely the authors’ responsibility and do not necessarily represent the official views of the funders (NHP, NINDS K23NS110980; KAL, AHA 26CDA1596118).

Acknowledgement statement

This work was supported by the NIH and the AHA (NHP, NINDS K23NS110980; KAL, AHA 26CDA1596118). KAL, LPT, DV, GJF, EJG, JK, JM-B, and RH report no relevant disclosures. AdH has received consultant fees from Novartis and Novo Nordisk and has equity in TitinKM and Certus. CCM serves as a consultant for Penumbra, Silk Road Medical, and MicroVention. SOG is a consultant for Medtronic, Stryker Neurovascular, and MicroVention with shares/stock options in BrainFlow, Motif, Gravity, and Eureka. NHP serves as a consultant for Silk Road Medical.

## AUTHOR CONTRIBUTIONS

KAL and NHP conceived and designed the study. Data acquisition, analysis, and interpretation were performed by KAL, NHP, CMM, and RH. Critical review of the manuscript for intellectual content, data analysis, and statistical design was performed by KAL, LPT, DV, GJF, EJG, JK, JM-B, AdH, KNS, CCM, SOG, RH, and NHP.

## DATA AVAILABILITY

The de-identified data and analysis routines that support the findings of this study are available from the corresponding author upon reasonable request, subject to institutional review and applicable data use agreements.

## SUPPORTING INFORMATION

The following supplementary material is available online:

**Supplementary File 1**.

**Table S1:** Demographic summaries for the derivation cohort of aneurysmal subarachnoid hemorrhage patients.

**Table S2:** Characteristics of the calibration and analysis LVO cohorts.

**Table S3:** Mixed-effects model coefficients predicting Scale_D_.

**Table S4:** Calibration model linear regression coefficients for predicting MAP_Opt_.

**Table S5:** Descriptive statistics for the LVO patients stratified by time within the predicted autoregulatory range.

**Table S6:** Distribution of 3-month modified Rankin Scale scores.

**Table S7:** Association between autoregulatory exposure and 3-month functional outcome.

**Table S8:** Partial proportional-odds model evaluating the association between time within the predicted autoregulatory range and 3-month mRS.

**Table S9:** Multivariable logistic regression predicting good functional outcome.

**Figure S1:** Overview of the analytic framework used to derive and apply an angiography-based estimate of cerebral autoregulatory state.

**Figure S2:** Angiography-derived perfusion processing pipeline for estimating mean transit time (MTT).

**Figure S3:** Model diagnostics for the final model including all observations.

**Figure S4:** Marginal effects of model predictors including all observations.

**Figure S5:** Model diagnostics after excluding an extreme observation.

**Figure S6:** Marginal effects after excluding the extreme observation.

**Figure S7:** Comparison of the primary linear calibration model with a 1-knot spline sensitivity model for angiography-to-NIRS delay.

**Figure S8:** Calibration performance of the angiography-derived model for predicting MAP_Opt_.

**Figure S9:** Bland-Altman analysis of angiography-derived MAP_Opt_ prediction.

**Figure S10:** Prediction accuracy of the angiography-derived MAP_Opt_ model across clinically relevant error thresholds.

**Figure S11:** Representative spatial maps of predicted MAP_Opt_ derived from Scale_D_ at time = 0.

**Figure S12:** Robustness of the association between time within the predicted autoregulatory range and 3-month functional outcome across alternative assumptions for the size of the range.

**Figure S13:** Sensitivity analysis comparing 3-month mRS distributions using the full Scale_D_-based model vs a reduced time-only model.

**Supplementary File 2**. STROBE Checklist

## References

1. Prabhakaran S, Gonzalez NR, Zachrison KS, et al. 2026 Guideline for the Early Management of Patients With Acute Ischemic Stroke: A Guideline From the American Heart Association/American Stroke Association. Stroke. Published online 2026. doi:10.1161/str.0000000000000513

2. Claassen JAHR, Thijssen DHJ, Panerai RB, Faraci FM. Regulation of cerebral blood flow in humans: physiology and clinical implications of autoregulation. Physiol Rev. 2021;101(4):1487–1559. doi:10.1152/physrev.00022.2020

3. Nogueira RC, Aries M, Minhas JS, et al. Review of studies on dynamic cerebral autoregulation in the acute phase of stroke and the relationship with clinical outcome. J Cereb Blood Flow Metab. 2021;42(3):430–453. doi:10.1177/0271678x211045222

4. Petersen NH, Begunova L, Olexa M, et al. Autoregulation-Guided Blood Pressure Targets After Stroke Thrombectomy: Impact on Secondary Brain Injury and Neurologic Outcomes. Neurology. 2026;106(3):e214577. doi:10.1212/wnl.0000000000214577

5. Petersen NH. Bedside Assessment of Cerebral Autoregulation: Working Toward a Common Monitoring Standard. Neurocritical Care. 2022;36(1):11–12. doi:10.1007/s12028-021-01304-2

6. Lyman KA, Rubin DB, Regenhardt RW, et al. Angiographic perfusion outperforms large artery vasospasm for predicting the impact of rescue therapy in subarachnoid hemorrhage. J Cereb Blood Flow Metab. Published online 2025:271678X251361992. doi:10.1177/0271678x251361992

7. Lyman KA, Hebert RM, Matouk CC, et al. Accounting for Truncation Artifacts in Angiographic Perfusion. Transl Stroke Res. 2026;17(2):32. doi:10.1007/s12975-026-01420-1

8. Petersen NH, Silverman A, Strander SM, et al. Fixed Compared With Autoregulation-Oriented Blood Pressure Thresholds After Mechanical Thrombectomy for Ischemic Stroke. Stroke. 2019;51(3):914–921. doi:10.1161/strokeaha.119.026596

9. Silverman A, Kodali S, Sheth KN, Petersen NH. Hemodynamics and Hemorrhagic Transformation After Endovascular Therapy for Ischemic Stroke. Front Neurol. 2020;11:728. doi:10.3389/fneur.2020.00728

10. Yang P, Song L, Zhang Y, et al. Intensive blood pressure control after endovascular thrombectomy for acute ischaemic stroke (ENCHANTED2/MT): a multicentre, open-label, blinded-endpoint, randomised controlled trial. Lancet. Published online 2022. doi:10.1016/s0140-6736(22)01882-7

11. Nam HS, Kim YD, Heo J, et al. Intensive vs Conventional Blood Pressure Lowering After Endovascular Thrombectomy in Acute Ischemic Stroke. JAMA. 2023;330(9):832–842. doi:10.1001/jama.2023.14590

