## Supplemental Materials for "Angiography-Derived Autoregulation Targets After Thrombectomy"

### Supplementary Material

#### 1. Additional Introduction information

Cerebral autoregulation refers to the capacity of the cerebral vasculature to maintain stable cerebral blood flow across a range of systemic arterial pressures.[1] Although the physiologic principles of autoregulation have been recognized for decades, clinical strategies to measure and maintain patients within individualized autoregulatory limits have not been widely adopted in routine practice. In prior work, we used continuous near-infrared spectroscopy (NIRS) monitoring to estimate patient-specific limits of autoregulation and demonstrated that the proportion of time spent within those limits was strongly associated with functional outcomes in patients with acute large-vessel occlusion (LVO).[2,3] These findings support the concept that deviations from a patient's autoregulatory range may represent a modifiable physiologic exposure relevant to neurologic recovery.

Despite this potential clinical importance, widespread implementation of autoregulation-guided management remains limited by the need for specialized monitoring equipment and continuous physiologic data acquisition.[4–6] Even under optimized research conditions, initiation of monitoring is often delayed, potentially missing a critical high-risk period after reperfusion. Recent work from our group has shown that quantitative perfusion metrics can be derived retrospectively from conventional digital subtraction angiography (DSA).[7,8]

In this study, we used angiography-derived perfusion features together with contemporaneous physiologic measurements to estimate a patient's position on the autoregulatory curve without reliance on continuous NIRS monitoring. We first derived the angiography-based autoregulatory metric ($Scale_D$) in an independent cohort of patients with aneurysmal subarachnoid hemorrhage (aSAH) who underwent simultaneous angiography and NIRS monitoring. We then calibrated this framework to predict optimal mean arterial pressure ($MAP_{Opt}$) in an LVO cohort and applied the resulting model to estimate time spent within predicted autoregulatory limits after thrombectomy. The analytic framework and perfusion processing pipeline are illustrated in Fig S1-2.

#### 2. Additional Methods information

##### 2.1 Derivation cohort (Aneurysmal Subarachnoid Hemorrhage, aSAH)

The derivation cohort consisted of patients 18 years or older who were admitted to the Neurosciences Intensive Care Unit (NICU) at Yale New Haven Hospital from September 2017 until March 2025 with aneurysmal subarachnoid hemorrhage. Eligible patients were identified through a prospectively maintained database and had been monitored with continuous near-infrared spectroscopy (NIRS) before and after digital subtraction angiography (DSA) during the admission as part of routine clinical care. The study was approved by the Yale Institutional Review Board with a waiver of individual informed consent due to its retrospective, observational design, and minimal risk to participants.

Of the eligible patients identified, only patients with at least one angiography run during DSA performed in the lateral view of sufficient quality to be analyzed were included (see below). Applying these criteria, we identified a cohort of 107 angiography runs performed from 28 patients on 50 distinct days. Baseline characteristics of the derivation cohort are summarized in Table S1.

2.2 Calibration and Analytic cohorts (Large Vessel Occlusion, LVO)

We conducted a secondary analysis of patients whose data had been prospectively collected in an observational study.[3] The parent study included 199 patients aged 18 years or older admitted to Yale New Haven Hospital between 2017 and 2022 with acute anterior circulation LVO who underwent endovascular thrombectomy. For this cohort, clinicians were blinded to the NIRS-derived autoregulation indices and hemodynamic management was independent of these measures.[3] Of these patients, 100 had angiography metadata stored in a format suitable for analysis and were eligible for inclusion. Fifteen patients were excluded because intraprocedural physiologic data were unavailable within the required 5-minute window (end-tidal carbon dioxide, n = 12; mean arterial pressure, n = 3). Two additional patients lacked post-thrombectomy lateral angiographic projections, leaving 85 patients. Of these, 64 had at least 5 seconds of analyzable angiographic run data after contrast bolus arrival. Two patients lacked 3-month modified Rankin Scale scores, resulting in a final cohort of 62 patients for the primary analysis. Baseline characteristics of the calibration and validation cohorts are summarized in Table S2. For the LVO calibration and analytic cohorts, the angiographic run used for MTT estimation was the final post-thrombectomy lateral ICA acquisition from the reperfused hemisphere; contralateral acquisitions were not required clinically and therefore were not available consistently enough for a paired contralateral analysis.

For calibration of the angiography-derived metric ScaleD in the context of large vessel occlusion, we initially restricted our calibration analysis to the 27 LVO patients whose angiography acquisitions lasted for at least 7 seconds after bolus arrival time (ScanTimeBAT $\geq$ 7 seconds, Fig S2B). This *a priori* threshold was chosen based on our prior results showing that 7 seconds or more of data after bolus arrival time is needed to estimate mean transit time with minimal error (<0.01 seconds error).[8] For the broader proof-of-concept clinical analysis, we relaxed the criteria for data inclusion to 5 seconds of data after bolus arrival time (ScanTimeBAT $\geq$5 seconds), which we previously associated with ~0.2 seconds of error in MTT estimates.[8] The more permissive 5-second threshold was used only for the proof-of-concept outcome analysis because underestimation of MTT from short acquisitions remains possible despite gamma-variate extrapolation and is treated as a limitation of the current implementation.

2.3 Angiography Processing Pipeline

All digital subtraction angiography (DSA) runs were analyzed using a previously validated computational pipeline for extracting quantitative perfusion metrics from conventional angiography.[7,8] Analyses were restricted to lateral projection images of the anterior circulation during contrast injection into the left or right internal carotid artery (ICA). For each angiography session, the corresponding DICOM files were retrieved, and individual angiographic runs were visually inspected for motion artifact. DSA runs lacking the metadata necessary to determine the acquisition timing of individual frames were excluded from analysis. The prior validation supported feasibility of deriving perfusion metrics from DSA and characterized truncation error, [7,8] but the present analysis did not include contemporaneous CT- or MR-perfusion validation of DSA-derived MTT in the LVO cohort.

All image processing was performed using custom-written MATLAB routines (R2025a, MathWorks, Natick, MA) as previously described, and briefly summarized here. To improve computational efficiency, all angiographic frames were spatially downsampled from the native size to 512 × 512 pixels. When variable frame rates were present (e.g., 4 Hz followed by 2 Hz or 1 Hz acquisition), the signal-time curves were temporally upsampled to a uniform sampling rate of 4 Hz. A rectangular region of interest (ROI) was manually placed over an extradural segment of the ICA. Signal intensity was converted to estimated contrast volume using cylindrical geometry and the average over time used to define the arterial input function (AIF) for the angiographic run. To isolate parenchymal pixels supplied by the ICA and minimize contamination from large vascular structures, a hemispheric ROI was manually drawn over the cortical territory of the lateral projection. Pixels representing large vascular structures were excluded by removing the 30% of pixels with the highest relative cerebral blood volume values. The bolus arrival time (BAT) was defined as the time point prior to the peak of the arterial input function where the second derivative of the AIF reached its maximum. The ScanTimeBAT metric was defined as the duration between the bolus arrival time and the end of the original angiographic acquisition as shown in Fig S2.[8] This parameter was used to assess whether sufficient imaging duration was available for reliable perfusion estimation.

To improve signal stability for curve fitting, each pixel's signal-time curve was spatially averaged with neighboring pixels within a 3-pixel radius, restricted to the remaining pixels contained within the hemispheric ROI[8]. Pixels identified as vascular structures were excluded from this spatial averaging step. To account for possible truncation of the signal-time curve and improve stability of perfusion estimates, the descending portion of each pixel's signal-time curve was modeled using a gamma-variate function[8]. Nonlinear least-squares fitting was performed using MATLAB's lsqcurvefit function with positivity constraints on the model parameters. Pixels were excluded from further analysis if the gamma-variate function failed to converge. Angiographic runs with less than twelve seconds of data after bolus arrival were extrapolated with the gamma-variate function to minimize the influence of truncation artifact.

Perfusion analysis was performed using block-circulant singular value decomposition (bcSVD) with a threshold of 0.2 after zero padding the signal to four times the length of the extrapolated signal-time curve, consistent with previously established perfusion imaging methods.[9] Relative cerebral blood volume (rCBV) was calculated as the integral of the deconvolved signal-time curve, and relative cerebral blood flow (rCBF) was defined as the maximum value of the residue function. Mean transit time (MTT) was then calculated according to the central volume theorem (MTT = rCBV/rCBF).[10]

2.4 Physiologic Data Extraction

Minute-by-minute physiologic data were extracted from the intraoperative anesthesia record, including end-tidal carbon dioxide ($EtCO_2$) and mean arterial pressure during angiography acquisition ($MAP_{Angio}$). These physiologic measurements were temporally aligned with digital subtraction angiography (DSA) acquisitions using timestamps contained in the DICOM metadata for each angiographic run. For each angiographic run, the physiologic exposure variables were defined as the mean $EtCO_2$ and mean MAP calculated across the 5-minute interval preceding and including the timestamp of the angiography acquisition. If multiple measurements were available within this interval, values were averaged. This timestamp-based linkage was used to ensure that the physiologic inputs reflected the same procedural period as the angiographic perfusion acquisition.

Quality control procedures were applied to remove missing or nonphysiologic measurements. $EtCO_2$ values were considered physiologically implausible and excluded if <10 mmHg. MAP measurements derived from arterial line recordings were excluded if the recorded systolic and diastolic pressures were identical or if the MAP was outside of the physiologic range (>200 mmHg or < 30 mmHg). When both invasive arterial pressure and noninvasive blood pressure measurements were recorded within the same interval, only the invasive arterial measurement was used. Angiography runs were included in the analysis if at least 1 valid measurement of both $EtCO_2$ and MAP could be made within the prespecified 5-minute window.

For patients in the large-vessel occlusion (LVO) cohort, the time of reperfusion was defined as the timestamp of the angiographic acquisition demonstrating reperfusion from DICOM metadata. If reperfusion was not achieved, the timestamp of the final angiographic run was used as the reference timepoint. Mean arterial pressure measurements during the 24-hour period following reperfusion were obtained from the electronic medical record and from physiologic data recorded using Intensive Care Monitor+ research software (ICM+, Cambridge Enterprise Ltd, Cambridge, United Kingdom), as previously described.[3] Physiologic traces were resampled to 1-minute resolution to generate a uniformly sampled time series. Short gaps in the raw data were linearly interpolated to maintain continuity of the MAP time series used for downstream analyses.

2.5 Definition of $Scale_D$

Our goal was to condense the information contained in the NIRS-derived autoregulatory curve into a single quantity that could be predicted from angiography and routine physiology (Fig 1A of the main text). We therefore defined $Scale_D$ as a normalized measure of the distance between MAP during angiography ($MAP_{Angio}$) and the patient's NIRS-derived $MAP_{Opt}$:

$$Scale_D = (MAP_{Angio} - MAP_{Opt}) / (ULA - LLA)$$

By construction, $Scale_D$ is positive when $MAP_{Angio}$ exceeds $MAP_{Opt}$ and negative when $MAP_{Angio}$ falls below $MAP_{Opt}$. Normalization by the width of the autoregulatory window (ULA - LLA) permits comparison across patients with differently sized autoregulatory ranges. Values near zero indicate pressure close to the estimated optimum, whereas larger positive or negative values indicate greater deviation above or below the optimum.

2.6 Model derivation, calibration, and operational definition of the predicted autoregulatory range

All statistical analyses were performed in SAS OnDemand for Academics (SAS Institute Inc, Cary, NC). In the derivation cohort of patients with aneurysmal subarachnoid hemorrhage (aSAH), we accounted for the potential influence of endovascular rescue therapy (ERT), as prior work from our group has demonstrated that ERT can alter angiography-derived perfusion metrics.[7] Accordingly, physiologic parameters used to compute $Scale_D$ were referenced to the timing of ERT relative to each angiographic acquisition. For each aSAH patient, $MAP_{Opt}$, the upper limit of autoregulation (ULA), and the lower limit of autoregulation (LLA) were calculated as the mean values observed within two predefined intervals: the 1 hour preceding and 1 hour following the time spent in the operating room for angiography. When an angiographic run occurred prior to ERT, the $Scale_D$ value for that run was calculated using $MAP_{Opt}$, ULA, and LLA derived from the 1-hour interval preceding angiography. When an angiographic run occurred after ERT, the ScaleD value was calculated using $MAP_{Opt}$, ULA, and LLA derived from the 1-hour interval following angiography**.** For vessels that did not undergo ERT, the $MAP_{Opt}$, ULA, and LLA estimates from the 1-hour pre-operating room and 1-hour post-operating room intervals were averaged, and these values were used to calculate $Scale_D$.

In the aSAH derivation cohort, $Scale_D$ was modeled using linear mixed-effects regression with a patient-level random intercept to account for repeated angiographic observations. Fixed effects included age, sex, MTT, $MAP_{Angio}$, $EtCO_2$, and prespecified interaction terms for MTT × $MAP_{Angio}$ and MTT × $EtCO_2$. Full coefficients for the derivation model are provided in Table S3. We then calibrated the angiography-derived $Scale_D$ metric to predict hemisphere-specific NIRS-derived $MAP_{Opt}$ in the LVO calibration subset using ordinary linear regression, with the mean of the first hour of NIRS-derived $MAP_{Opt}$ values as the dependent variable and $Scale_D$ plus minutes elapsed from reperfusion until the first available NIRS-derived $MAP_{Opt}$ measurement as predictors. Regression coefficients for this calibration step are shown in Table S4. As a sensitivity analysis, we also examined a 1-knot spline model for elapsed time, with the knot placed at the median time point of the 27-patient calibration cohort (562 minutes after reperfusion).

For the proof-of-concept clinical application, predicted autoregulatory limits were defined operationally as predicted $MAP_{Opt}$ ± 20 mmHg. Sensitivity analyses repeated the outcome models using alternative operational ranges of predicted $MAP_{Opt}$ ± 10 mmHg and ± 30 mmHg.

2.7 Outcome analysis

The primary clinical outcome was 3-month modified Rankin Scale (mRS), treated as a 7-level ordinal variable (0–6). Functional outcomes were determined through a telephone interview at 3 months by a member of the research team who was blinded to monitoring results.[3] The primary exposure was the percentage of time during the first 24 hours after reperfusion during which minute-by-minute MAP remained within the predicted autoregulatory range. The primary multivariable model was a proportional-odds ordinal logistic regression adjusted for age, sex, baseline mRS, admission National Institutes of Health Stroke Scale score, Alberta Stroke Program Early CT Score, intravenous thrombolysis, and reperfusion status. We assessed the proportional-odds assumption and, when it was not fully satisfied, performed a sensitivity analysis using an adjusted cumulative-logit partial proportional-odds model in which the time-within-range exposure was allowed threshold-specific effects. For clinical interpretability, we also fit an adjusted binary logistic regression for good functional outcome (mRS 0–2). Reperfusion status was included in the clinical outcome models to account for differences in procedural success. It was not included in the $Scale_D$ derivation or calibration model because those models were designed to translate angiographic perfusion and contemporaneous physiology into an autoregulatory estimate; the sample size was not sufficient to fit separate calibration models by occlusion location or incomplete-reperfusion subgroup.

**3. Additional Results information**

The supplemental results are organized into three sections: (3.1) derivation of the angiography-based autoregulatory metric ($Scale_D$) in the aneurysmal subarachnoid hemorrhage cohort, (3.2) calibration of the $MAP_{Opt}$ prediction model in the calibration subset of the large vessel occlusion (LVO) cohort, and (3.3) application of the resulting framework to the full LVO analysis cohort to evaluate associations between predicted autoregulatory exposure and functional outcome.

3.1 Development of $Scale_D$ in the aSAH derivation cohort

The derivation cohort included 107 angiographic runs from 28 patients with aSAH (Table S1). The linear mixed-effects model identified significant associations between $Scale_D$ and $EtCO_2$, MTT, and $MAP_{Angio}$, together with significant interaction terms indicating that the relationship between pressure, carbon dioxide, and autoregulatory position depended on the underlying perfusion state (Table S3). Age and sex were not independently associated with $Scale_D$ after multivariable adjustment.

Across the full dataset, observed and model-predicted $Scale_D$ values were strongly correlated, with residuals centered near zero and only modest right-tail distortion caused by a single extreme observation (Fig S3). Marginal-effect plots in the full dataset showed that the direction and magnitude of the relationship between $MAP_{Angio}$ and $Scale_D$ varied across the observed range of MTT and similarly that the association between $EtCO_2$ and $Scale_D$ depended on transit time (Fig S4).

Because a single observation broadened the visual range of the diagnostic plots, we repeated the graphical presentation after excluding that point from the visualization alone. These restricted plots showed similar overall behavior, including residual symmetry and preservation of the principal interaction patterns, supporting model stability across the clustered range containing nearly all observations (Fig S5 and Fig S6).

3.2: Calibration of the angiography-derived predictor in LVO

Clinical and physiologic characteristics of the LVO calibration subset and the full angiographic cohort are shown in Table S2. In the 27-patient calibration subset with ScanTimeBAT $\geq$ 7 seconds, $Scale_D$ and elapsed time from angiography to NIRS jointly predicted first-hour NIRS-derived $MAP_{Opt}$ (Table S4). Within this higher-quality calibration subset, the model explained 41% of the variance in $MAP_{Opt}$, with root mean squared error of 9.6 mmHg and mean absolute error of 7.3 mmHg.

When the calibrated model was applied across the 64-patient LVO cohort, predicted $MAP_{Opt}$ showed minimal overall bias with moderate patient-level precision (Fig S8). Residuals were centered near zero, and inspection of the residual-versus-predicted and residual-versus-time plots suggested some remaining nonlinear structure.

As a sensitivity analysis for the calibration step, we examined a 1-knot spline as an alternative calibration regression, placing the knot at the median time point of the 27-patient calibration cohort (562 minutes after reperfusion). This choice provided a stable, prespecified bend near the center of the observed training distribution while avoiding overfitting in the small sample. The 1-knot spline did not materially improve the predictive accuracy compared to the linear regression (Fig S7). The mean absolute error (MAE) was 8.042 for the 1-knot spline vs 8.051 for the linear regression. Given the limited number of observations in the calibration cohort, more elaborate models were not considered, and the linear regression was used for simplicity.

Bland-Altman analysis showed a mean bias of −0.28 mmHg, with 95% limits of agreement from −20.82 to 20.25 mmHg (Fig S9). Accuracy analyses showed that 43.8% of predictions fell within ±5 mmHg, 68.8% within ±10 mmHg, and 84.4% within ±15 mmHg of observed $MAP_{Opt}$ (Fig S10). Based on these calibration results, the primary clinical analyses used predicted $MAP_{Opt}$ ± 20 mmHg as the operational autoregulatory range, with sensitivity analyses at ±10 mmHg and ±30 mmHg. Taken together, these findings suggest that the current model should be interpreted primarily as a physiology-informed estimator rather than a point-precise surrogate for NIRS-derived $MAP_{Opt}$.

To illustrate the spatial information retained by the angiography-based approach, we also generated pixel-wise maps of predicted $MAP_{Opt}$ by applying the $Scale_D$-based model to MTT at each analyzable pixel. Representative examples are shown in Fig S11 and demonstrate regional heterogeneity in predicted autoregulatory targets that is not captured by a single global summary value.

3.3 Functional outcomes after thrombectomy

Among the 64 patients with analyzable angiography, 32 spent more time within the predicted autoregulatory range and 32 spent less time within that range during the first 24 hours after reperfusion. Descriptive characteristics for these groups are provided in Table S5. The distribution of 3-month mRS outcomes in the 62 patients with available follow-up is shown in Table S6.

In the primary adjusted proportional-odds model, greater time within the predicted autoregulatory range was independently associated with better 3-month functional outcome (odds ratio per 10% increase, 1.86; 95% CI, 1.31-2.66; P = .0006) (Table S7). In a partial proportional-odds sensitivity analysis, the common proportional component for time within range remained significant (odds ratio, 1.57; 95% CI, 1.01-2.47; P = .048), whereas the global unequal-slope component was not significant (Wald $\chi^2$ = 7.87; P = .16) (Table S8). In adjusted binary logistic regression, greater time within the predicted autoregulatory range was also associated with higher odds of good functional outcome (mRS 0–2) (odds ratio, 3.92; 95% CI, 1.44-10.68; P = .0075) (Table S9). Across alternative definitions of the autoregulatory range, the association remained significant for predicted $MAP_{Opt}$ ±10 mmHg, ±20 mmHg, and ±30 mmHg (Fig S12).

To test whether the physiology-derived $Scale_D$ component added meaningful information beyond elapsed time alone, we repeated the workflow using a reduced time-only calibration model. This approach omitted $Scale_D$ from the linear calibration model (Fig S1B) and instead calibrated predicted $MAP_{Opt}$ as a linear function of time ($MAP_{Opt} = \alpha_0 + Time^* \alpha_1$). The full model separated 3-month outcome distributions more clearly than the reduced model, supporting the incremental value of the physiology-derived term (Fig S13).

Because contralateral angiography was not consistently available, we did not compare ipsilateral and contralateral predicted $MAP_{Opt}$. Similarly, the cohort was underpowered for formal subgroup testing by occlusion location or by individual reperfusion-score category; reperfusion status was therefore handled as an adjustment covariate in the outcome models rather than as a stratification variable in the calibration model.

**4. Additional Discussion information**

In this supplemental analysis, three findings support the main report. First, angiography-derived perfusion features and contemporaneous physiologic variables predicted a NIRS-derived estimate of autoregulatory position in the aSAH derivation cohort. Second, that metric could be calibrated to $MAP_{Opt}$ in the LVO cohort with minimal overall bias but limited individual-level precision. Third, despite imperfect patient-level calibration, greater time within the predicted autoregulatory range after thrombectomy was consistently associated with better 3-month functional outcome across proportional-odds, partial proportional-odds, binary logistic, and autoregulation-modeling sensitivity analyses. These findings suggest that routinely acquired angiography and procedural physiology contain clinically relevant information about patient-specific autoregulatory state.

The LVO cohort analyzed here represents a subset of patients included in our prior work using near-infrared spectroscopy (NIRS)–derived estimates of optimal mean arterial pressure ($MAP_{Opt}$)[3]. Notably, the magnitude of association observed in the present analysis was larger than that previously reported, with each 10% increase in time spent within predicted autoregulatory limits associated with a 1.86-fold increase in the odds of favorable functional outcome. This difference may reflect the earlier physiologic window captured here. However, because the angiography-based framework provided less distinction between patient groups than NIRS and likely narrowed the dynamic range of the exposure metric, these findings should be interpreted as supportive rather than directly comparable.

Despite only moderate patient-level agreement between predicted and NIRS-derived $MAP_{Opt}$, the association between time within the predicted autoregulatory range and functional outcome remained robust. The primary analysis used predicted $MAP_{Opt}$ ± 20 mmHg as the operational range, and sensitivity analyses using ±10 mmHg and ±30 mmHg yielded similar results (Fig S12). These findings suggest that the clinically relevant signal may lie less in point-precise estimation of $MAP_{Opt}$ than in identifying whether patients spend substantial time meaningfully above or below a physiology-informed pressure target. The true optimal pressure target for improving outcome remains uncertain, and NIRS itself provides only an imperfect, spatially limited estimate of autoregulatory state. Future work incorporating region-specific tissue risk, potentially using spatial $MAP_{Opt}$ maps (Fig S11), may help refine hemodynamic target selection.

Unlike NIRS-based approaches, which require prolonged monitoring before stable $MAP_{Opt}$ estimates can be derived, the present model enables estimation of autoregulatory targets immediately following reperfusion. In the cohort examined here, the first NIRS-derived $MAP_{Opt}$ estimate occurred a mean of 708 minutes after reperfusion (Table S2), effectively excluding the early post-reperfusion period from prior analyses. This interval may represent a particularly vulnerable phase, during which fluctuations in arterial pressure could exert disproportionate effects on microvascular perfusion and tissue recovery.[12,13] Consistent with this hypothesis, our prior work demonstrated progressive increases in time spent within autoregulatory limits over time, suggesting that autoregulatory stability improves in the hours following reperfusion.[2,3] Analyses that begin later in the clinical course may therefore underestimate the physiologic importance of early autoregulatory exposure.

Across multiple modeling frameworks including proportional odds, partial proportional odds, and binary logistic regression, the association between time spent within predicted autoregulatory limits and functional outcome remained robust. These findings suggest that early autoregulatory exposure is not only measurable using angiography-derived features, but also strongly linked to neurologic recovery. By enabling estimation of autoregulatory targets at the time of angiography, this framework provides a practical approach for studying, and potentially guiding, blood pressure management during a critical window of cerebral vulnerability.

At the same time, the current framework highlights several opportunities to improve patient-level accuracy. The present implementation reduces spatially resolved perfusion maps to a single summary measure of mean transit time, potentially obscuring regional heterogeneity in autoregulatory state (Fig S11). Future approaches that incorporate territory-specific perfusion features may better capture physiologic variation within the ischemic brain. Similarly, although a linear relationship between time and $MAP_{Opt}$ was assumed for model calibration, the true temporal evolution of autoregulatory targets is likely more complex and may require larger datasets with continuous physiologic monitoring to characterize.[4]

Several limitations should be considered. First, prediction of autoregulatory limits relied on a simplified operational definition centered on $MAP_{Opt}$, which does not capture potential asymmetry or dynamic changes in autoregulatory range. Second, the observational design precludes determination of whether deviations from predicted autoregulatory targets are causally related to neurologic injury or instead reflect underlying disease severity. Despite these limitations, the present findings support the fundamental premise that angiography-derived physiologic features contain quantifiable information about patient-specific autoregulatory state. By enabling estimation of autoregulatory targets immediately following angiography without specialized monitoring hardware, this approach offers a scalable and clinically accessible framework for investigating autoregulation in acute neurovascular disease. Additional limitations include the absence of contemporaneous CT- or MR-perfusion validation, derivation of the $Scale_D$ model in an aSAH cohort with different pathophysiology from LVO, reliance on NIRS-derived $MAP_{Opt}$ as the training reference, and the lack of systematic contralateral angiography for side-to-side comparison.

Future work should focus on improving predictive accuracy through incorporation of additional physiologic variables, exploration of nonlinear modeling strategies, and validation in larger, prospectively collected datasets. Modest optimization of angiographic acquisition, including slightly longer runs, may improve the fidelity with which perfusion angiography captures cerebral hemodynamics. In addition, dedicated acquisitions performed at distinct blood pressures could allow estimation of an individual patient's autoregulatory range rather than reliance on population-based thresholds, creating a potential pathway toward intraoperative personalization of blood pressure targets. Ultimately, prospective interventional studies will be necessary to determine whether angiography-derived, autoregulation-guided blood pressure management can improve neurologic outcomes following reperfusion therapy.

**Figure S1. Overview of the analytic framework used to derive and apply an angiography-based estimate of cerebral autoregulatory state.**
**Panel A (Model Development).** In the derivation cohort of patients with aneurysmal subarachnoid hemorrhage (aSAH), angiography-derived perfusion metrics and physiologic variables were incorporated into a linear mixed-effects model to estimate a patient's position on the NIRS-derived autoregulatory curve ($Scale_D$). The model incorporated age, sex, mean transit time (MTT), mean arterial pressure during angiography ($MAP_{Angio}$), end-tidal carbon dioxide ($etCO_2$), and interaction terms.
**Panel B (Model Calibration).** In a calibration subset of patients with large-vessel occlusion (LVO) and high-quality angiography acquisitions (ScanTimeBAT ≥7 seconds), the derived $Scale_D$ parameter was combined with elapsed time from angiography to predict the NIRS-derived optimal mean arterial pressure ($MAP_{Opt}$) using linear regression.
**Panel C (Clinical Application).** The calibrated model was then applied to the full LVO analysis cohort to estimate $MAP_{Opt}$ and compute the proportion of time spent within predicted limits of autoregulation during the 24-hour period following reperfusion, without reliance on NIRS monitoring.
Abbreviations: aSAH, aneurysmal subarachnoid hemorrhage; LVO, large-vessel occlusion; $MAP_{Angio}$, mean arterial pressure measured during angiography; $MAP_{Opt}$, optimal mean arterial pressure; MTT, mean transit time; NIRS, near-infrared spectroscopy.

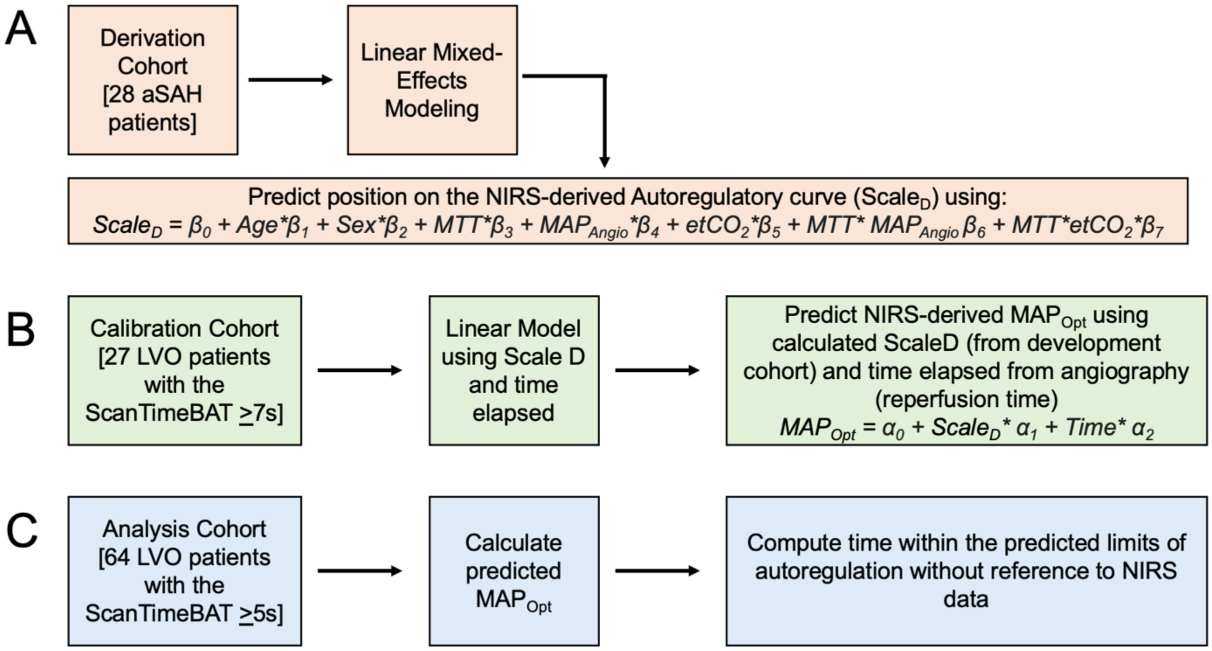

**Figure S2. Angiography-derived perfusion processing pipeline for estimating mean transit time (MTT).**
Schematic overview of the workflow used to extract perfusion parameters from digital subtraction angiography (DSA) and derive physiologic features for downstream modeling.
**Panel A (Image preprocessing).** High-quality angiographic runs obtained in the lateral projection were selected and standardized through spatial downsampling and temporal resampling to ensure consistent resolution across acquisitions. A rectangular region of interest (ROI) was placed within the extradural internal carotid artery to define the arterial input function (AIF), while a hemispheric cortical ROI was used to capture tissue-level signal.
**Panel B (Bolus arrival time).** The AIF was used to define bolus arrival time (BAT).
**Panel C (Analysis mask).** The ‘Raw DSA Image’ panel shows a representative arterial phase image highlighting vascular structures. The middle image, ‘-Vascular Pixels’, displays in white the remaining pixels after removing large vascular structures using the relative cerebral blood volume (rCBV). After spatially averaging the remaining pixels, gamma-variate fitting was applied to the signal-time curve for each of the remaining pixels and those that failed to converge were removed. The ‘-Pixels without fitting’ image shows the remaining pixels where the gamma-variate function converged.
**Panel D (Mean Transit Time).** The ‘Unfiltered Map’ shows the result of calculating MTT for every available pixel from the DSA. After removing the pixels using the vascular and gamma-variate filters (**Panel C**), the remaining pixels are shown in the ‘Analysis Map’. Averaging the remaining pixels for this representative angiography run yielded an MTT of 3.69 seconds. Abbreviations: AIF, arterial input function; BAT, bolus arrival time; DSA, digital subtraction angiography; MTT, mean transit time; rCBV, relative cerebral blood volume; ROI, region of interest.

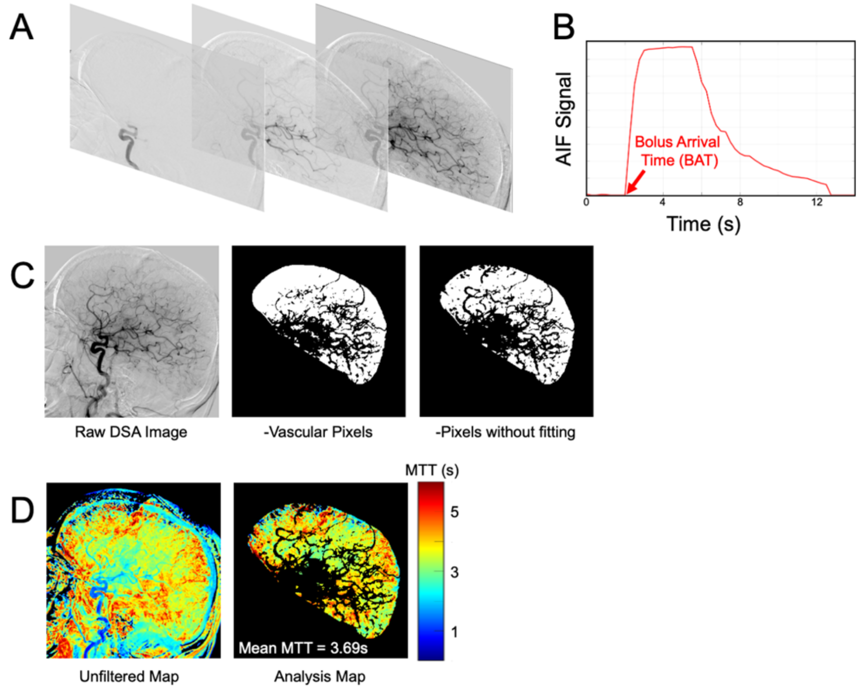

**Figure S3. Model Diagnostics for the Final Model Including All Observations**
**A**, Observed versus predicted values for $Scale_D$. Each point represents an individual observation. The dashed line indicates perfect agreement (y = x), and the solid line represents the linear fit between observed and predicted values. Summary performance metrics are shown in the panel (N = 107 observations; Pearson r = 0.833; root mean squared error [RMSE] = 0.731; mean absolute error [MAE] = 0.554; bias = 0.051).
**B**, Residual versus fitted values for the full dataset. The dashed horizontal line indicates zero residual error. The smoothed curve illustrates the local trend of residuals across the range of predicted values.
**C**, Distribution of residuals for the full dataset with kernel density overlay, demonstrating approximate symmetry around zero with a small right-sided tail influenced by a single extreme observation.

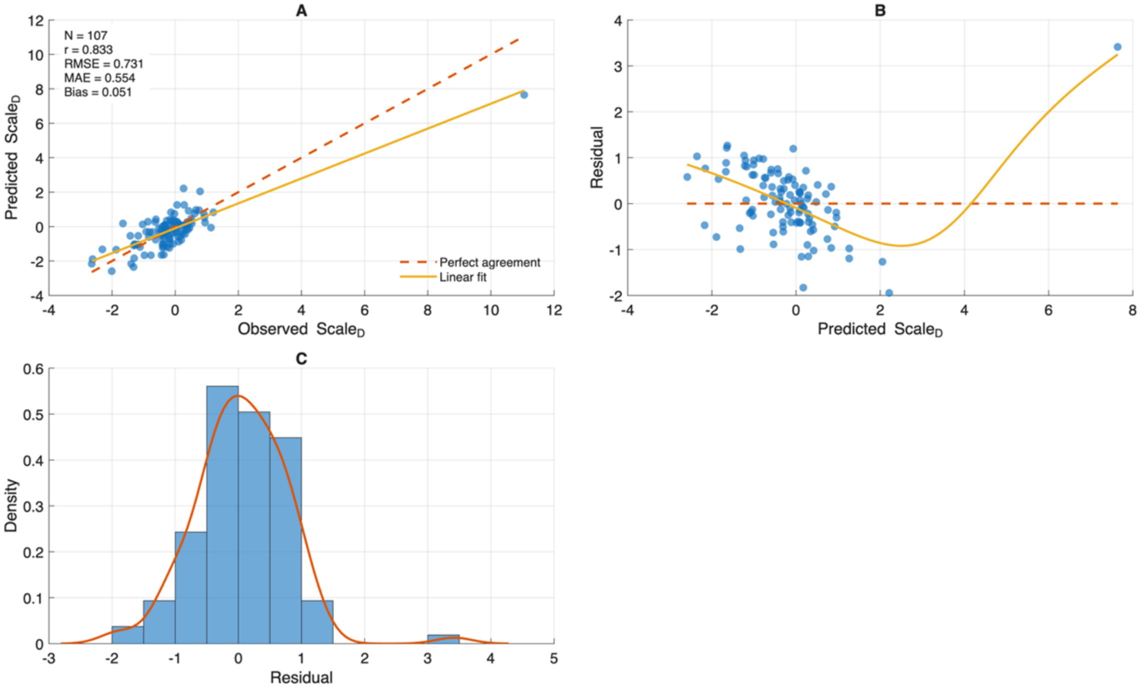

**Figure S4. Marginal Effects of Model Predictors Including All Observations**
Predicted values of the $Scale_D$ outcome across the observed range of mean transit time (MTT).
**A**, Interaction between MTT and mean arterial pressure measured during angiography ($MAP_{Angio}$). Predicted values are shown for representative $MAP_{Angio}$ values corresponding to approximately the 25th, 50th, and 75th percentiles of the dataset while holding other covariates constant ($etCO_2$ = 30 mmHg, age = 63 years, male sex).
**B**, Interaction between MTT and end-tidal carbon dioxide ($etCO_2$). Predicted values are shown for representative $etCO_2$ values corresponding to approximately the 25th, 50th, and 75th percentiles while holding other covariates constant ($MAP_{Angio}$ ≈ 105 mmHg, age = 63 years, male sex).

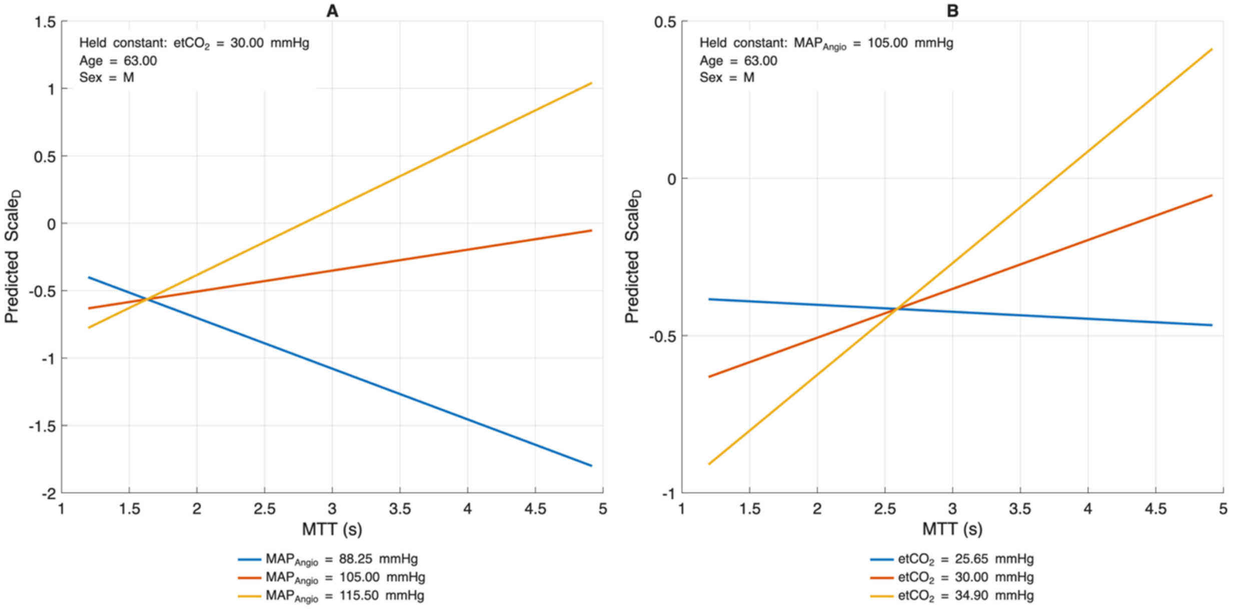

**Figure S5. Model Diagnostics After Excluding an Extreme Observation**
Diagnostic plots restricted to observations with $Scale_D \leq 10$ (N = 106 observations).
**A**, Observed versus predicted values demonstrating model calibration within the clustered range of the dataset. Performance metrics are shown (Pearson r = 0.671; RMSE = 0.655; MAE = 0.527; bias = 0.020).
**B**, Residual versus fitted values showing the distribution of residuals across predicted values without the extreme observation.
**C**, Histogram of residuals with kernel density overlay demonstrating approximately symmetric residual distribution centered near zero.
These plots are provided to facilitate visualization of model behavior within the range containing most of the observations.

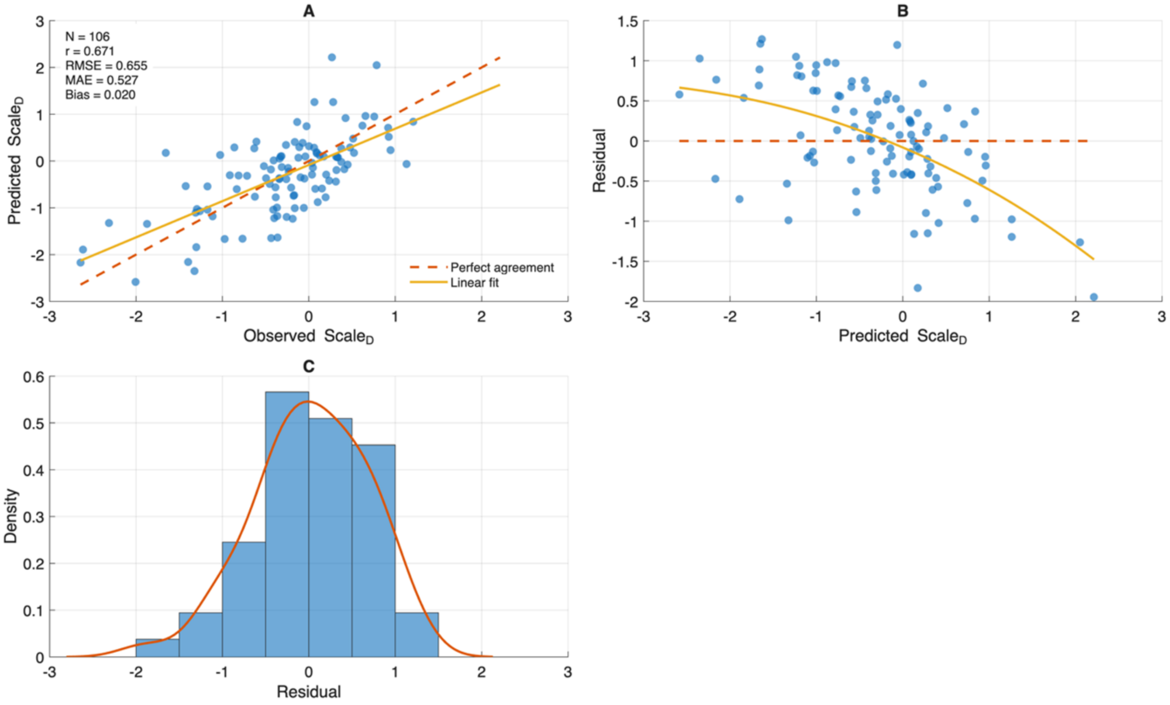

**Figure S6. Marginal Effects After Excluding the Extreme Observation**
Marginal effect plots restricted to observations with $Scale_D \leq 10$.
**A**, Interaction between MTT and $MAP_{Angio}$ across representative values of $MAP_{Angio}$ (88, 105, and 115 mmHg).
**B**, Interaction between MTT and $etCO_2$ across representative values of $etCO_2$ (26, 30, and 35 mmHg).
Predicted values are shown while holding remaining covariates constant (age = 63 years; male sex).

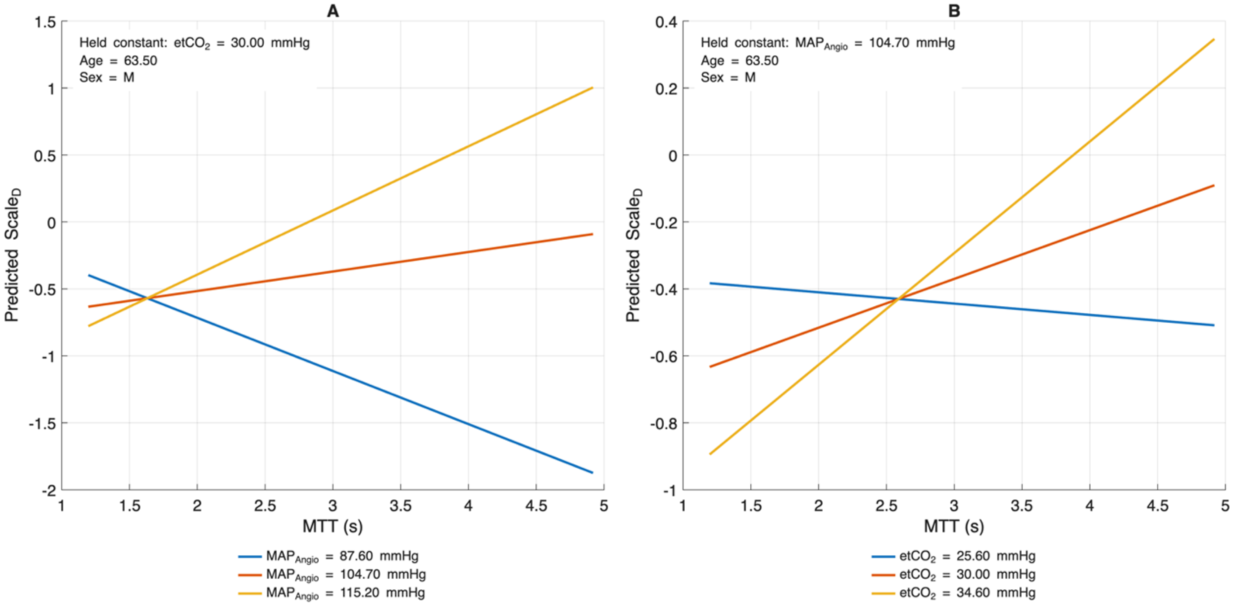

**Figure S7. Comparison of the primary linear calibration model with a 1-knot spline sensitivity model for angiography-to-NIRS delay.**
**Panel A**, In the calibration cohort (n=27), predicted $MAP_{Opt}$ is shown as a function of time from reperfusion until the first NIRS-derived $MAP_{Opt}$ for the primary linear model and a 1-knot spline model with the knot placed at the training-cohort median delay (562 minutes). Curves are displayed with $Scale_D$ fixed at its cohort mean to isolate the relationship between delay and predicted $MAP_{Opt}$.
**Panel B**, Per-observation absolute error for the 1-knot spline model is plotted against absolute error for the linear model in the analytic large-vessel occlusion cohort (n=64). The diagonal line denotes equality of error between models.
**Panel C**, Absolute error for the linear model across the analytic large-vessel occlusion cohort, with the dashed horizontal line indicating the cohort mean absolute error (MAE).
**Panel D,** Absolute error for the 1-knot spline model across the same cohort, with the solid horizontal line indicating the cohort MAE. The spline model yielded only trivial changes in predictive performance relative to the linear model. In the analytic large-vessel occlusion cohort, MAE was nearly identical (8.051 vs 8.042) and $R^2$ was slightly lower for the spline model (Linear: 0.195 vs Spline: 0.182), supporting use of the simpler linear specification as the primary analysis.

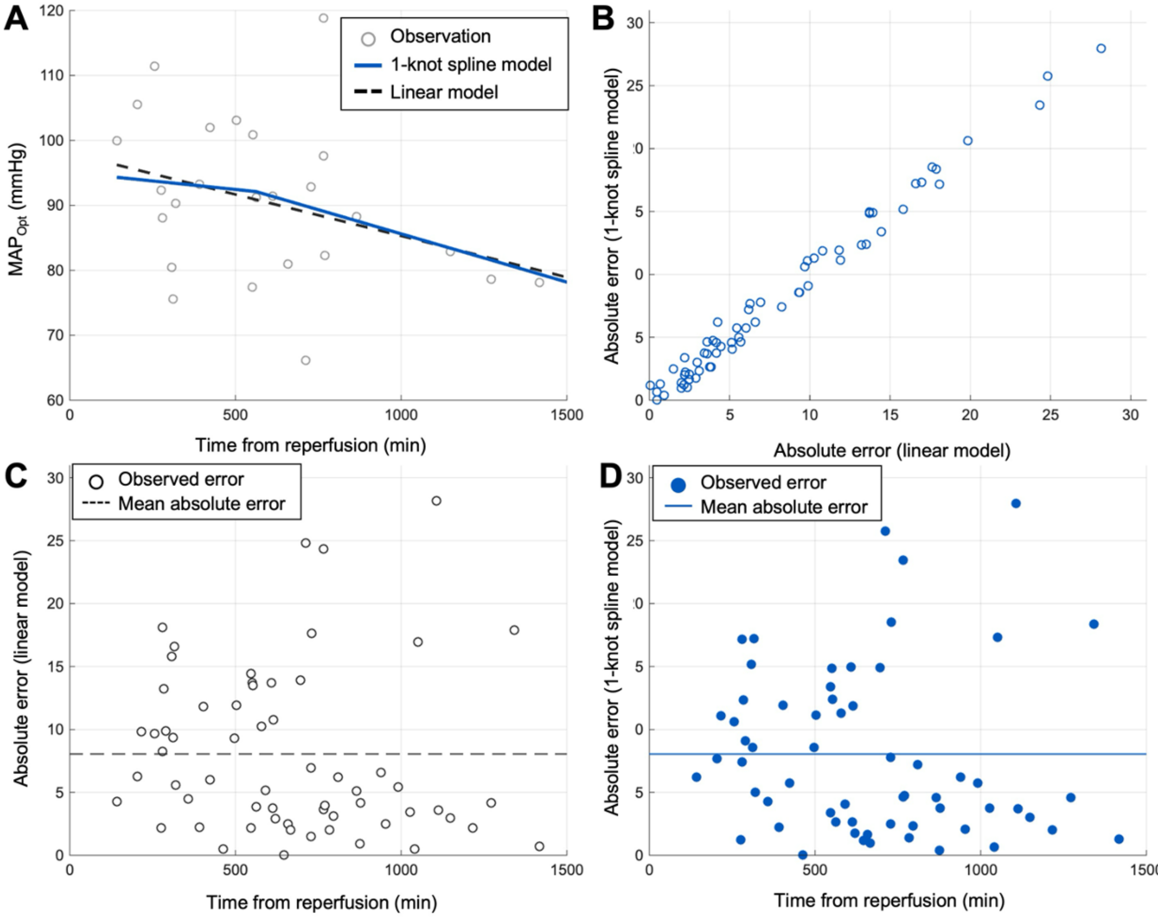

**Figure S8. Calibration performance of the angiography-derived model for predicting $MAP_{Opt}$.** Diagnostic plots for the fixed model predicting observed $MAP_{Opt}$ using the physiologic predictor $Scale_D$ and minutes elapsed from angiography until NIRS in the analysis cohort (N=64). In all residual plots, residuals are defined as observed $MAP_{Opt}$ − predicted $MAP_{Opt}$, so positive values indicate underprediction by the model and negative values indicate overprediction.
**Panel A, observed vs predicted $MAP_{Opt}$:** Each point represents 1 patient-level observation. The dashed diagonal line denotes perfect agreement (predicted = observed). The solid line denotes the least-squares linear fit to the scatter (Intercept = 62.79 mmHg, slope = 0.29). Model-level summary statistics are shown within the panel: Pearson correlation r = 0.470; RMSE = 10.397 mmHg; MAE = 8.051 mmHg; and mean error (bias) = −0.285 mmHg, indicating minimal systematic over- or underestimation overall.
**Panel B, residual vs predicted $MAP_{Opt}$:** Residuals are plotted against fitted values to evaluate calibration across the predicted range and to assess model misspecification. The horizontal dashed line marks zero residual. The solid smoothed line represents a locally estimated scatterplot smoothing (LOESS) trend, included to visualize any systematic departure from zero across fitted $MAP_{Opt}$ values.
**Panel C, residual distribution:** Histogram of residuals with an overlaid kernel density estimate (solid curve), illustrating the empirical distribution of prediction errors. Residuals were centered near zero (mean = −0.285 mmHg; median = 0.215 mmHg), with interquartile range from −6.564 to 4.695 mmHg (IQR, 11.259 mmHg).
**Panel D, normal Q-Q plot of residuals:** Ordered residuals are plotted against expected normal quantiles. The dashed reference line indicates the theoretical relationship expected under approximate normality. Deviation from this line, particularly in the tails, reflects departure from a strictly Gaussian error distribution.
**Panel E, residual vs Minutes Elapsed from Angiography to NIRS:** Residuals are plotted against elapsed time to assess whether time-related bias remains after inclusion of time in the model. The horizontal dashed line marks zero residual; the solid curve represents a LOESS smoother.
**Panel F, predicted $MAP_{Opt}$ vs Minutes Elapsed from Angiography to NIRS:** Predicted $MAP_{Opt}$ is plotted against elapsed time to visualize the modeled time dependence. The solid curve is a LOESS smoother summarizing the empiric relationship between elapsed time and model-predicted $MAP_{Opt}$.

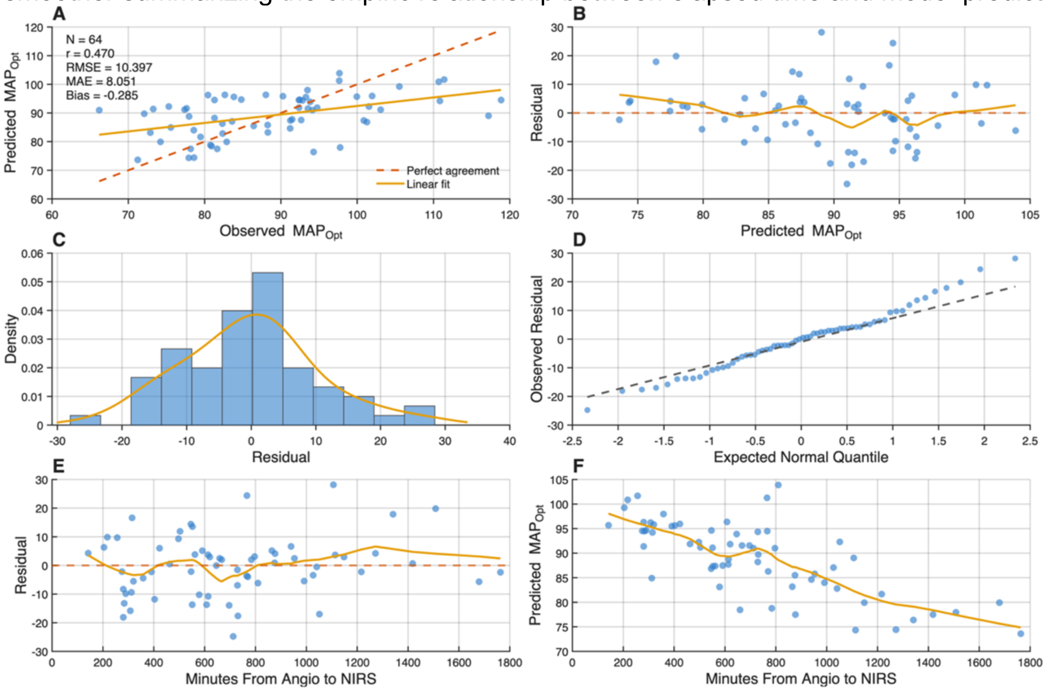

**Figure S9. Bland-Altman Analysis of Angiography-Derived $MAP_{Opt}$ Prediction**
Bland-Altman plot comparing observed NIRS-derived $MAP_{Opt}$ and model-predicted $MAP_{Opt}$ across the analytic cohort. The x-axis shows the mean of observed and predicted $MAP_{Opt}$ for each patient, and the y-axis shows the difference between observed and predicted $MAP_{Opt}$ (observed − predicted). The solid horizontal line denotes the mean bias, and the dashed horizontal lines denote the 95% limits of agreement (mean bias = −0.28 mmHg, 95% limits of agreement −20.82 to 20.25 mmHg). The mean bias was small, indicating limited systematic overestimation or underestimation at the cohort level, although the width of the limits of agreement demonstrates substantial patient-level prediction error. $MAP_{Opt}$ indicates optimal mean arterial pressure.

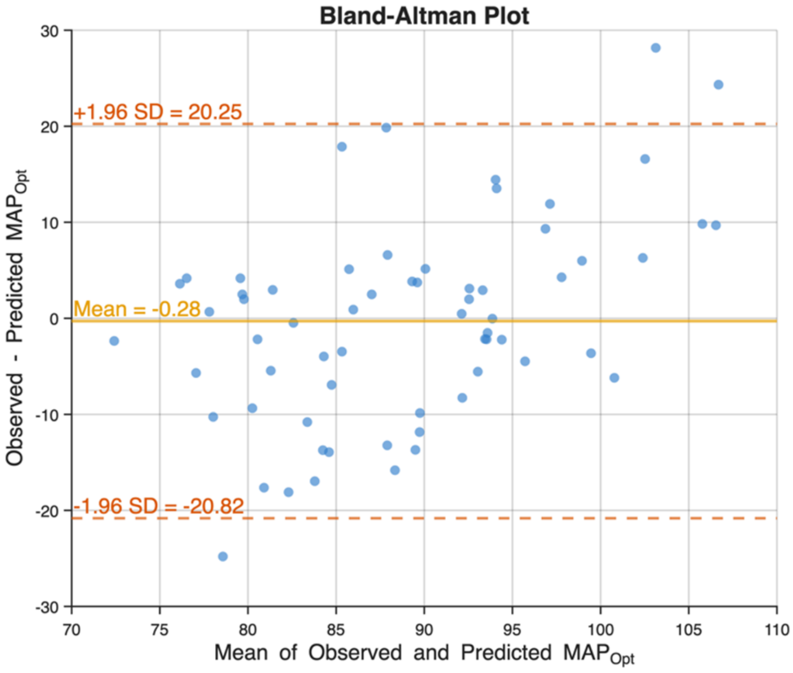

**Figure S10. Prediction Accuracy of the Angiography-Derived $MAP_{Opt}$ Model Across Clinically Relevant Error Thresholds**

Bar plot showing the proportion of model predictions that fell within prespecified absolute error thresholds relative to observed $MAP_{Opt}$. Percentages are shown for predictions within ±5 mm Hg, ±10 mm Hg, and ±15 mm Hg of the observed value. Accuracy increased across wider error bands, with approximately 43.8% of predictions within ±5 mm Hg, 68.8% within ±10 mm Hg, and 84.4% within ±15 mm Hg, demonstrating moderate agreement at narrow thresholds and good agreement at clinically relevant ranges. $MAP_{Opt}$ indicates optimal mean arterial pressure.

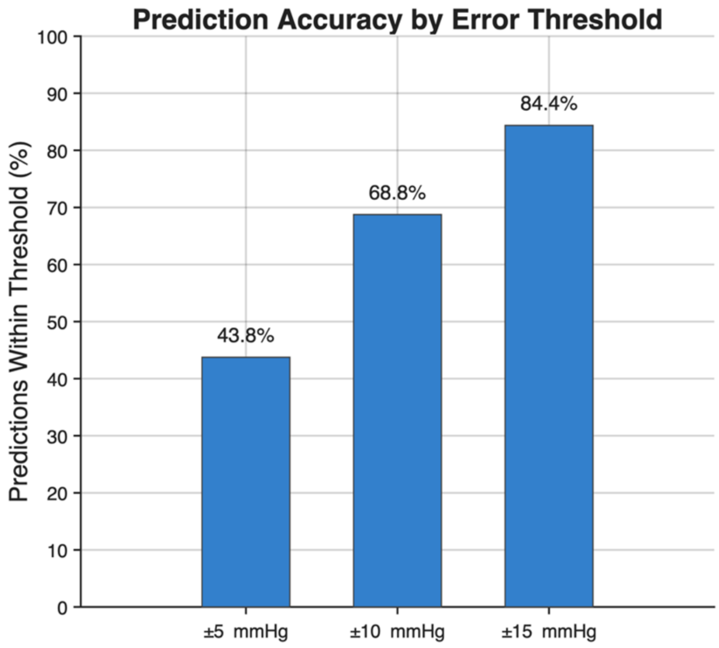

**Figure S11. Representative spatial maps of predicted $MAP_{Opt}$ derived from $Scale_D$ at time = 0.**
Predicted optimal mean arterial pressure ($MAP_{Opt}$) was calculated for each angiographic pixel by applying the $Scale_D$-based predictive model to the mean transit time (MTT) measured at that pixel. Rather than averaging MTT values across the entire region of interest prior to prediction, $MAP_{Opt}$ was computed on a pixel-by-pixel basis, generating spatial maps that illustrate regional heterogeneity in predicted autoregulatory targets.
**Patient #1** shows an example from an 87-year-old woman with end-tidal $CO_2$ of 36.7 mmHg, mean transit time 1.97 s, $Scale_D$ = −0.81, and intra-angiographic mean arterial pressure ($MAP_{Angio}$) of 95.3 mmHg.
**Patient #2** shows a second example from an 88-year-old man with end-tidal $CO_2$ 20.0 mmHg, mean transit time 3.72 s, $Scale_D$ = −1.81, and $MAP_{Angio}$ 92.7 mmHg.
These examples illustrate how the predictive pipeline can generate spatially resolved estimates of $MAP_{Opt}$, allowing visualization of regional physiologic variation across the angiographic field rather than producing a single patient-level estimate and highlighting regional heterogeneity in autoregulatory targets that is not captured by global summary measures.

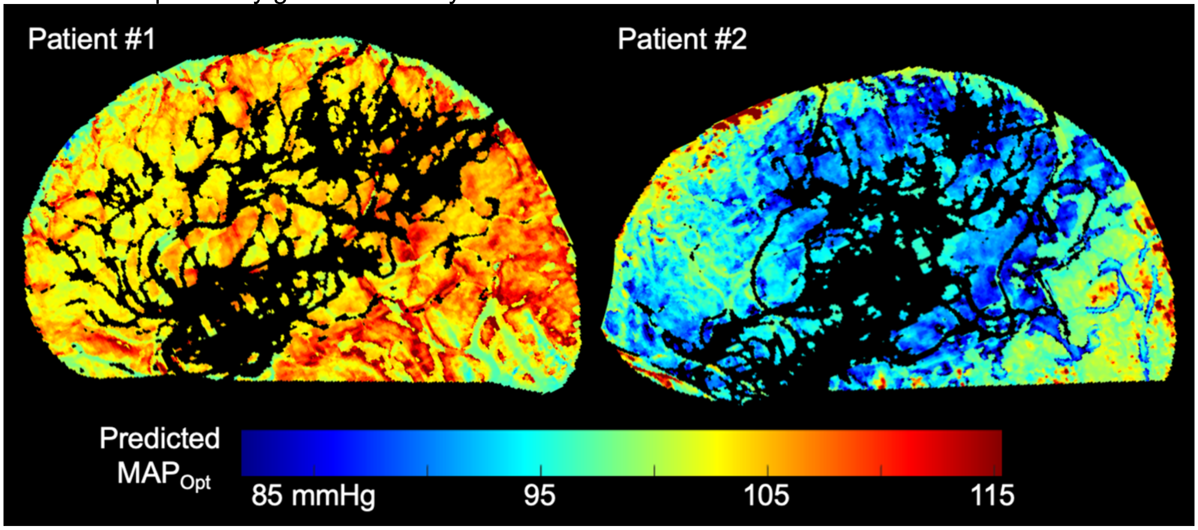

**Figure S12. Robustness of the association between time within the predicted autoregulatory range and 3-month functional outcome across alternative assumptions for the size of the range**
**Top row (Ai-Ci),** representative post-thrombectomy mean arterial pressure (MAP) trajectory from a single patient, shown against the predicted $MAP_{Opt}$ and alternative operational limits defined as predicted $MAP_{Opt}$ ± 10 mmHg, ± 20 mmHg, and ± 30 mmHg. As the bandwidth widens, a greater proportion of the same MAP trajectory is classified as within the predicted autoregulatory range.
**Bottom row (Aii-Cii),** adjusted ordinal logistic forest plots for 3-month modified Rankin Scale (mRS) in the 62-patient analytic cohort, with the primary exposure defined as the percentage of time during the first 24 hours after reperfusion that minute-by-minute MAP remained within each alternative range. Models were adjusted for age, sex, baseline mRS, admission National Institutes of Health Stroke Scale score, Alberta Stroke Program Early CT Score, intravenous thrombolysis, and angiographic reperfusion grade (recoded on an ordinal scale). Across all three bandwidth definitions, greater time within the predicted autoregulatory range remained independently associated with a favorable shift in 3-month mRS. For the ±10 mmHg definition, the odds ratio per 10% increase in time within range was 1.58 (95% CI, 1.19-2.11; P = .0017); for the ±20 mmHg definition, 1.86 (95% CI, 1.31-2.66; P = .0006); and for the ±30 mmHg definition, 2.57 (95% CI, 1.33-4.97; P = .0049). Model discrimination was similar across bandwidths (c statistics, 0.810, 0.814, and 0.791, respectively), supporting a directionally consistent and statistically robust association despite varying the operational definition of the predicted autoregulatory range. The covariate pattern was also stable across models. Intravenous thrombolysis, older age, higher admission NIHSS score, and greater baseline disability remained significantly associated with outcome at each bandwidth. Reperfusion score (recoded on an ordinal scale) remained significant for the ± 10 mmHg and ± 20 mmHg models but was attenuated at ± 30 mmHg (OR, 1.49; 95% CI, 0.97-2.29; P = .07), whereas sex and ASPECTS were not independently associated with outcome in any of the three models shown.

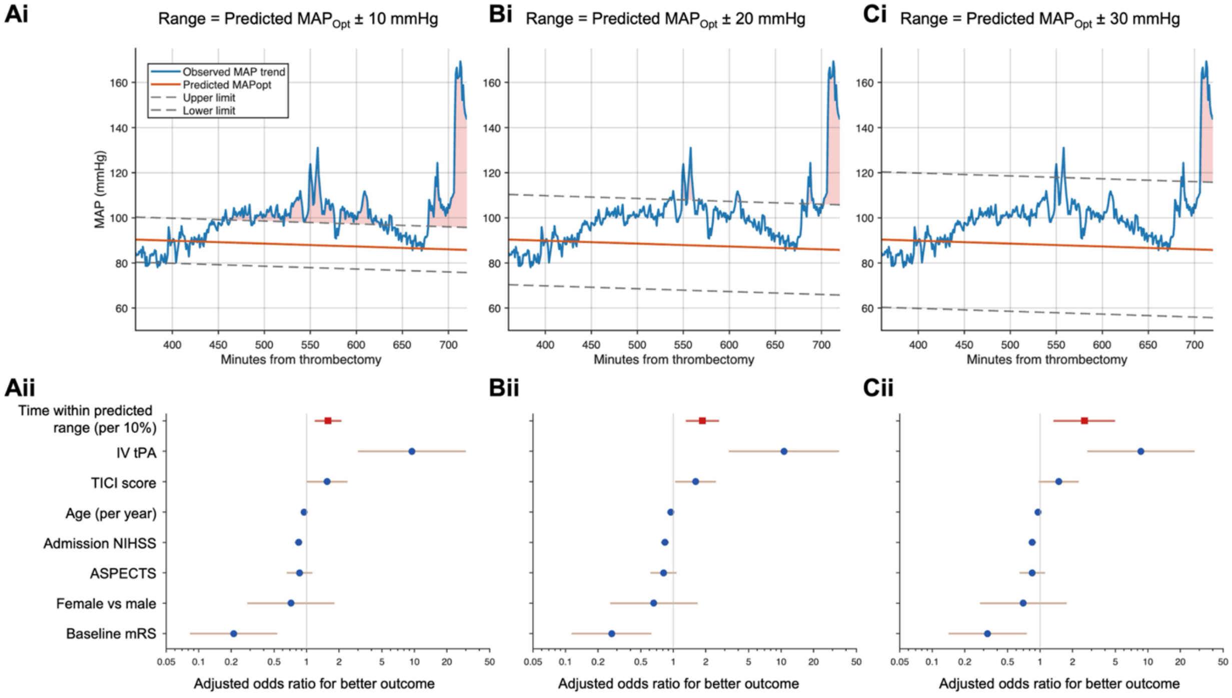

**Figure S13. Sensitivity analysis comparing 3-month mRS distributions using the full $Scale_D$-based model vs a reduced time-only model.**
Horizontal stacked bars show the distribution of 3-month modified Rankin Scale (mRS) scores among patients stratified by time spent within each model's predicted limits of autoregulation, defined as predicted $MAP_{Opt}$ ± 20 mmHg during the 24 hours after angiography.
**Panel A** shows results for the full model, in which predicted $MAP_{Opt}$ was calibrated using both the physiology-derived parameter $Scale_D$ and elapsed time from angiography.
**Panel B** shows results for the reduced model, in which predicted $MAP_{Opt}$ was calibrated using elapsed time alone, without $Scale_D$. Within each panel, patients were dichotomized into groups spending more vs less time within the model-predicted autoregulatory range (n=31 per group). The heavy vertical line marks the boundary between mRS 0-2 and mRS 3-6, with the diagonal connector illustrating the shift in the cumulative distribution between groups. The full model demonstrated separation in 3-month functional outcome distributions, whereas the time-only model did not, indicating that inclusion of $Scale_D$ is necessary to predict clinically meaningful outcomes (3-month mRS). mRS, modified Rankin Scale.

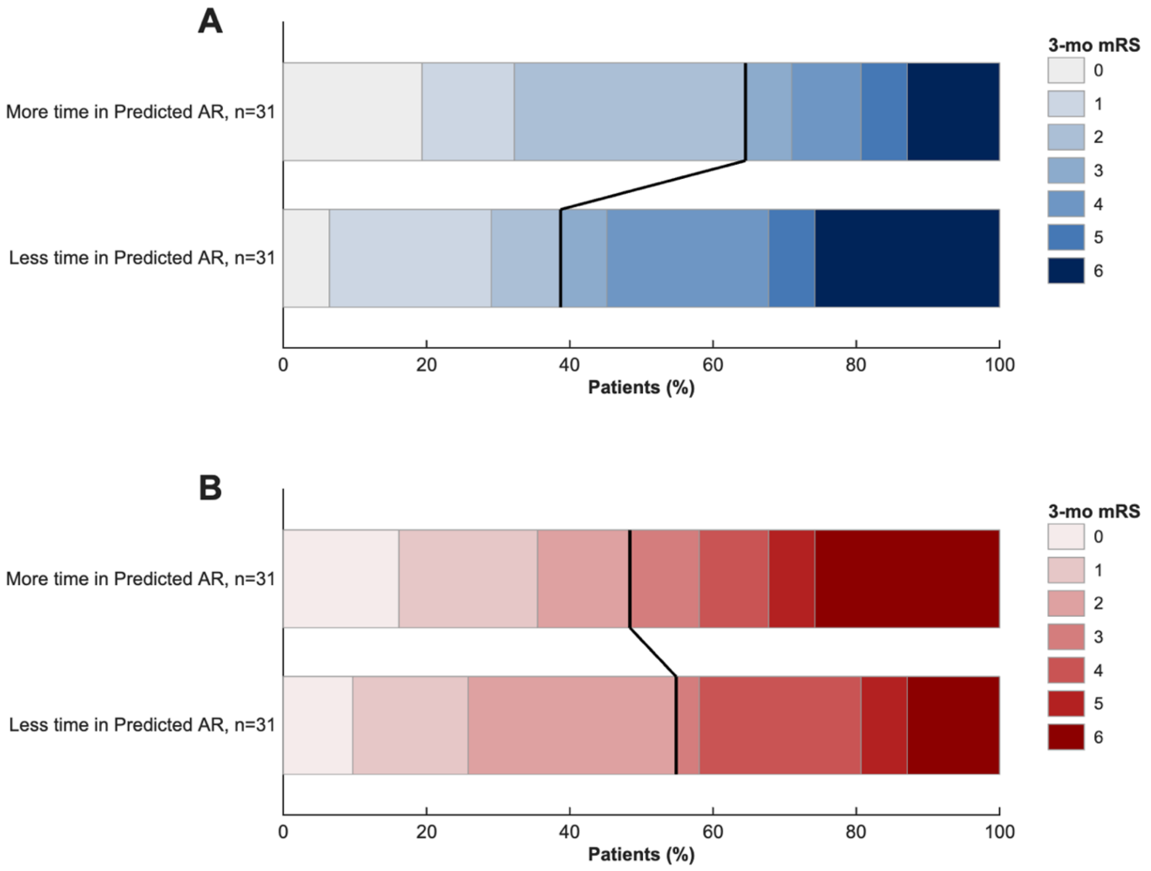

**Table S1, Demographic summaries for the derivation cohort of aneurysmal subarachnoid hemorrhage patients.**
Demographic and clinical characteristics of the aneurysmal subarachnoid hemorrhage (aSAH) cohort used for model derivation. Top portion displays the patient-level data and the bottom portion displays observation-level data. Continuous variables are presented as mean (SD) or median (IQR) as appropriate.

| **Characteristic** | **Value** |
|---|---|
| **Demographic and clinical characteristics (per patient)** | |
| Age, years | 58.8 (12.4) |
| Female sex, No. (%) | 16 (57.1) |
| Male sex, No. (%) | 12 (42.9) |
| Hunt and Hess grade, median [IQR] | 3.5 [3-4] |
| Modified Fisher grade, median [IQR] | 4 [4-4] |
| **Physiologic characteristics (per angiography run)** | |
| Days after rupture, mean (SD) | 7.4 (3.4) |
| $MAP_{Angio}$, mean (SD), mmHg | 103.9 (19.9) |
| $EtCO_2$, mean (SD), mmHg | 28.9 (7.5) |
| MTT, mean (SD), s | 3.08 (0.74) |
| $MAP_{Opt}$ (NIRS), mean (SD), mmHg | 109.9 (14.2) |
| ULA (NIRS), mean (SD), mmHg | 125.0 (14.6) |
| LLA (NIRS), mean (SD), mmHg | 94.5 (13.7) |
| $MAP_{Angio}$ - $MAP_{Opt}$, mean (SD), mmHg | -5.9 (21.8) |
| ULA - LLA (NIRS), mean (SD), mmHg | 30.4 (9.7) |
| $Scale_D$, mean (SD) | -0.17 (1.32) |

**Abbreviations:** aSAH, aneurysmal subarachnoid hemorrhage; EtCO2, end-tidal carbon dioxide; LLA, lower limit of autoregulation; MAPAngio, mean arterial pressure during angiography; MAPopt, optimal mean arterial pressure; MTT, mean transit time; NIRS, near-infrared spectroscopy; ScaleD, position on the NIRS-derived autoregulatory curve; ULA, upper limit of autoregulation.

**Table S2, Characteristics of the Calibration and Analysis LVO Cohorts**
Demographic, clinical, and physiologic characteristics of patients with large-vessel occlusion used in the calibration and analysis cohorts. Parentheses denote standard deviation and brackets denote interquartile range. Note that the calibration cohort is a subset of the full analysis cohort. The TICI variable is reported as the ordinal reperfusion-score recoded on an ordinal scale rather than as a literal modified TICI grade so that higher values indicate more complete reperfusion.

| Characteristic | Calibration Cohort | Full Cohort |
|---|---|---|
| Patients, No. | 27 (subset of full cohort) | 64 |
| Age, mean (SD), years | 69.5 (17.8) | 70.5 (14.8) |
| Female sex, No. (%) | 10 (37%) | 31 (48.4%) |
| $EtCO_2$, mean (SD), mmHg | 25.1 (7.0) | 26.9 (8.2) |
| $Scale_D$, mean (SD) | -0.58 (0.83) | -0.56 (0.81) |
| Time within predicted autoregulatory range, mean (SD), % | 80.2 (17.7) | 82.9 (15.4) |
| MTT, mean (SD), s | 2.99 (0.71) | 2.91 (0.99) |
| $MAP_{Angio}$, mean (SD), mmHg | 101.8 (16.1) | 102.5 (16.2) |
| Admission NIHSS, median [IQR] | 15 [8-19] | 14.5 [7.5-19] |
| ASPECTS, median [IQR] | 7.5 [7-10] | 8 [7-10] |
| IV thrombolysis, No. (%) | 13 (48.1) | 28 (43.7) |
| TICI score, median [IQR] | 5 [3-5] | 4 [3-5] |
| Time from reperfusion until NIRS, mean (SD), min | 675.4 (438.5) | 708.5 (369.9) |

**Abbreviations:** ASPECTS, Alberta Stroke Program Early CT Score; EtCO2, end-tidal carbon dioxide; MAPAngio, mean arterial pressure during angiography; MTT, mean transit time; NIHSS, National Institutes of Health Stroke Scale; NIRS, near-infrared spectroscopy; ScaleD, position on the NIRS-derived autoregulatory curve; TICI, Thrombolysis in Cerebral Infarction. TICI, ordinal recode of reperfusion status used for analysis rather than a literal mTICI category.

**Table S3, Mixed-Effects Model Coefficients Predicting $Scale_D$**
Fixed-effect coefficients, confidence intervals, and statistical significance for the linear mixed-effects model used to estimate $Scale_D$, including interaction terms demonstrating effect modification between physiologic variables.

| Predictor | β coefficient | 95% CI | P value |
|---|---|---|---|
| Main effects | | | |
| $EtCO_2$ | -0.106 | -0.189 to -0.022 | 0.014 |
| MTT | -4.40 | -5.81 to -2.99 | <0.001 |
| $MAP_{Angio}$ | -0.052 | -0.085 to -0.019 | 0.003 |
| Age, years | -0.011 | -0.032 to 0.010 | 0.29 |
| Female sex (vs male) | 0.26 | -0.27 to 0.78 | 0.32 |
| Interaction terms | | | |
| MTT x $MAP_{Angio}$ | 0.031 | 0.022 to 0.041 | <0.001 |
| MTT x $EtCO_2$ | 0.040 | 0.015 to 0.066 | 0.002 |

**Abbreviations:** CI, confidence interval; EtCO2, end-tidal carbon dioxide; MAPAngio, mean arterial pressure during angiography; MTT, mean transit time.

**Table S4, Calibration model linear regression coefficients for predicting $MAP_{Opt}$**

A linear regression model predicting $MAP_{Opt}$ from $Scale_D$ and elapsed time from angiography to NIRS was trained in patients with ≥7 seconds of data after angiographic bolus arrival time (n=27). The resulting model was predicted $MAP_{Opt} = \alpha_0 + \alpha_1 * (Scale_D) + \alpha_2 * (Time)$. When evaluated within the same physiologic range (ScanTimeBAT ≥7), the model explained 41% of the variance in $MAP_{Opt}$ ($R^2 = 0.41$) with a root mean squared error of 9.6 mmHg and mean absolute error of 7.3 mmHg.

| Parameter | Value |
|---|---|
| α0 | 101.65 |
| α1 | 6.13 |
| α2 | -0.012 |

**Abbreviations:** $MAP_{Opt}$, optimal mean arterial pressure; ScanTimeBAT, length of the original data acquisition after bolus arrival time, expressed in seconds.

**Table S5, Descriptive statistics for the LVO patients stratified by time within the predicted autoregulatory range.**

The analyzable cohort of patients with large-vessel occlusion (n=64) was divided into the 50% who spent more time in the predicted autoregulatory range and the 50% who spent less time within the predicted autoregulatory range. Mean values are presented with standard deviation in parentheses while median values are shown with interquartile range in brackets.

| Characteristic | More Time in Predicted Autoregulatory Range | Less Time in Predicted Autoregulatory Range |
|---|---|---|
| Patients, No. | 32 | 32 |
| Age, mean (SD), years | 71.9 (13.9) | 69.2 (15.7) |
| Female sex, No. (%) | 14 (43.8) | 17 (53.1) |
| $EtCO_2$, mean (SD), mmHg | 26.9 (7.6) | 26.9 (8.9) |
| $Scale_D$, mean (SD) | -0.80 (0.66) | -0.32 (0.88) |
| Time within predicted autoregulatory range, median [IQR], % | 94.4 [91.9-97.1] | 75.4 [62.1-84.4] |
| MTT, mean (SD), s | 2.98 (1.03) | 2.84 (0.96) |
| $MAP_{Angio}$, mean (SD), mmHg | 100.3 (17.4) | 104.6 (14.9) |
| Admission NIHSS, median [IQR] | 14.5 [6-19] | 14 [8-19] |
| ASPECTS, median [IQR] | 8 [6.75-10] | 8 [7-9.5] |
| IV thrombolysis, No. (%) | 13 (40.6) | 15 (46.9) |
| TICI score, median [IQR] | 4 [3-5] | 4 [3-5] |

**Abbreviations:** ASPECTS, Alberta Stroke Program Early CT Score; EtCO2, end-tidal carbon dioxide; MAPAngio, mean arterial pressure during angiography; MTT, mean transit time; NIHSS, National Institutes of Health Stroke Scale; ScaleD, position on the NIRS-derived autoregulatory curve; TICI, Thrombolysis in Cerebral Infarction. TICI, ordinal recode of reperfusion status used for analysis rather than a literal mTICI category.

**Table S6, Distribution of 3-Month Modified Rankin Scale Scores**
Distribution of functional outcomes at 3 months in the LVO analysis cohort.

| Outcome | No. (%) |
|---|---|
| **Modified Rankin Scale score** | |
| 0 | 8 (12.9) |
| 1 | 11 (17.7) |
| 2 | 13 (21.0) |
| 3 | 4 (6.5) |
| 4 | 10 (16.1) |
| 5 | 4 (6.5) |
| 6 | 12 (19.4) |
| **Dichotomized functional outcome** | |
| Good outcome (mRS 0-2) | 32 (51.6) |
| Poor outcome (mRS 3-6) | 30 (48.4) |

**Abbreviations:** mRS, modified Rankin Scale.

**Table S7, Association between autoregulatory exposure and 3-month functional outcome**
Results of the multivariable ordinal logistic regression evaluating the relationship between time spent within predicted autoregulatory limits and 3-month modified Rankin Scale outcome. Time within the predicted autoregulatory range is scaled per 10% increase in time within the predicted autoregulatory limits and odds ratios greater than 1 indicate a favorable shift toward lower Modified Rankin Scale score.

| Predictor | Odds ratio | 95% CI | P value |
|---|---|---|---|
| Time within predicted autoregulatory range (10%) | 1.86 | 1.31-2.66 | 0.0006 |
| Age, per year | 0.95 | 0.92-0.99 | 0.007 |
| Sex (female vs male) | 0.66 | 0.26-1.68 | 0.38 |
| IV tPA | 10.63 | 3.28-34.51 | <0.001 |
| Baseline mRS | 0.27 | 0.11-0.63 | 0.002 |
| Admission NIHSS | 0.83 | 0.77-0.91 | <0.001 |
| TICI score | 1.61 | 1.04-2.49 | 0.033 |
| ASPECTS | 0.81 | 0.61-1.07 | 0.14 |

**Abbreviations:** ASPECTS, Alberta Stroke Program Early CT Score; CI, confidence interval; mRS, modified Rankin Scale; NIHSS, National Institutes of Health Stroke Scale; TICI, Thrombolysis in Cerebral Infarction; tPA, tissue plasminogen activator.

**Table S8. Partial proportional-odds model evaluating the association between time within the predicted autoregulatory range and 3-month mRS.**
Partial proportional-odds cumulative-logit model for 3-month mRS, with probabilities cumulated over lower ordered values. The proportion of time within the predicted autoregulatory range was the only predictor allowed nonproportional, threshold-specific effects; all other covariates were constrained to have proportional effects. The common proportional component represents the average association of the proportion of time within the predicted autoregulatory range across mRS cut points, whereas the unequal-slope component tests departure from proportional odds.

| Predictor | Parameterization | OR | 95% CI | Wald $\chi^2$ | P value |
|---|---|---|---|---|---|
| Time within predicted autoregulatory range | Common proportional component, per 10% increase | 1.57 | 1.01-2.47 | 3.93 | 0.048 |
| Time within predicted autoregulatory range | Global unequal-slope component | NA | NA | 7.87 | 0.16 |
| Age | Per year | 0.95 | 0.92-0.99 | 7.54 | 0.006 |
| Sex | Female vs male | 0.66 | 0.25-1.73 | 0.70 | 0.40 |
| IV tPA | Yes vs no | 10.53 | 3.11-35.71 | 14.28 | 0.0002 |
| Baseline mRS | Per point | 0.25 | 0.10-0.62 | 8.82 | 0.003 |
| Admission NIHSS | Per point | 0.84 | 0.77-0.91 | 16.02 | <0.001 |
| TICI score | Per category (ordinal) | 1.62 | 1.06-2.46 | 5.06 | 0.025 |
| ASPECTS | Per point | 0.82 | 0.62-1.08 | 2.05 | 0.15 |

**Threshold-specific deviation terms for the proportion of time within the predicted autoregulatory range**

| mRS threshold | Estimate (SE) | P value |
|---|---|---|
| mRS ≤0 vs >0 | 0.185 (0.428) | 0.66 |
| mRS ≤1 vs >1 | -0.180 (0.393) | 0.64 |
| mRS ≤2 vs >2 | 0.582 (0.406) | 0.15 |
| mRS ≤3 vs >3 | 0.581 (0.312) | 0.06 |
| mRS ≤4 vs >4 | 0.089 (0.160) | 0.57 |
| mRS ≤5 vs >5 | Reference | NA |

**Note:** NA indicates not applicable.
**Abbreviations:** ASPECTS, Alberta Stroke Program Early CT Score; CI, confidence interval; mRS, modified Rankin Scale; NIHSS, National Institutes of Health Stroke Scale; OR, odds ratio; SE, standard error; TICI, Thrombolysis in Cerebral Infarction; tPA, tissue plasminogen activator.

**Table S9, Multivariable Logistic Regression Predicting Good Functional Outcome**
Adjusted logistic regression model evaluating predictors of good functional outcome (mRS 0–2). Time within the predicted autoregulatory range is scaled to reflect a 10% increase in time.

| Predictor | OR | 95% CI | P value |
|---|---|---|---|
| Time within predicted autoregulatory range (10%) | 3.92 | 1.44-10.68 | 0.0075 |
| Age, per year | 0.93 | 0.86-1.00 | 0.050 |
| Female sex | 1.26 | 0.26-6.08 | 0.77 |
| IV tPA | 26.11 | 2.99-227.77 | 0.003 |
| Baseline mRS | 0.36 | 0.10-1.27 | 0.11 |
| Admission NIHSS | 0.76 | 0.64-0.90 | 0.001 |
| TICI score | 1.26 | 0.66-2.41 | 0.52 |
| ASPECTS | 0.66 | 0.41-1.05 | 0.082 |

**Abbreviations:** ASPECTS, Alberta Stroke Program Early CT Score; CI, confidence interval; mRS, modified Rankin Scale; NIHSS, National Institutes of Health Stroke Scale; OR, odds ratio; TICI, Thrombolysis in Cerebral Infarction; tPA, tissue plasminogen activator.

**SUPPLEMENTAL REFERENCES**

1. Claassen JAHR, Thijssen DHJ, Panerai RB, Faraci FM. Regulation of cerebral blood flow in humans: physiology and clinical implications of autoregulation. *Physiol Rev*. 2021;101(4):1487-1559. doi:10.1152/physrev.00022.2020

2. Petersen NH, Silverman A, Wang A, et al. Association of Personalized Blood Pressure Targets With Hemorrhagic Transformation and Functional Outcome After Endovascular Stroke Therapy. *JAMA Neurol*. 2019;76(10):1256-1258. doi:10.1001/jamaneurol.2019.2120

3. Petersen NH, Begunova L, Olexa M, et al. Autoregulation-Guided Blood Pressure Targets After Stroke Thrombectomy: Impact on Secondary Brain Injury and Neurologic Outcomes. *Neurology*. 2026;106(3):e214577. doi:10.1212/wnl.0000000000214577

4. Budohoski KP, Czosnyka M, Kirkpatrick PJ, Smielewski P, Steiner LA, Pickard JD. Clinical relevance of cerebral autoregulation following subarachnoid haemorrhage. *Nat Rev Neurol*. 2013;9(3):152-163. doi:10.1038/nrneurol.2013.11

5. Wang A, Ortega-Gutierrez S, Petersen NH. Autoregulation in the Neuro ICU. *Curr Treat Options Neurol*. 2018;20(6):20. doi:10.1007/s11940-018-0501-x

6. Petersen NH. Bedside Assessment of Cerebral Autoregulation: Working Toward a Common Monitoring Standard. *Neurocritical Care*. 2022;36(1):11-12. doi:10.1007/s12028-021-01304-2

7. Lyman KA, Rubin DB, Regenhardt RW, et al. Angiographic perfusion outperforms large artery vasospasm for predicting the impact of rescue therapy in subarachnoid hemorrhage. *J Cereb Blood Flow Metab*. Published online 2025:271678X251361992. doi:10.1177/0271678x251361992

8. Lyman KA, Hebert RM, Matouk CC, et al. Accounting for Truncation Artifacts in Angiographic Perfusion. *Transl Stroke Res*. 2026;17(2):32. doi:10.1007/s12975-026-01420-1

9. Wu O, Østergaard L, Weisskoff RM, Benner T, Rosen BR, Sorensen AG. Tracer arrival timing-insensitive technique for estimating flow in MR perfusion-weighted imaging using singular value decomposition with a block-circulant deconvolution matrix. *Magn Reson Med*. 2003;50(1):164-174. doi:10.1002/mrm.10522

10. Wu O, Østergaard L, Sorensen AG. Technical Aspects of Perfusion-Weighted Imaging. *Neuroimaging Clin North Am*. 2005;15(3):623-637. doi:10.1016/j.nic.2005.08.009

11. Bath PMW, Lees KR, Schellinger PD, et al. Statistical Analysis of the Primary Outcome in Acute *Stroke* Trials. Stroke. 2012;43(4):1171-1178. doi:10.1161/strokeaha.111.641456

12. Sheriff F, Castro P, Kozberg M, et al. Dynamic Cerebral Autoregulation Post Endovascular Thrombectomy in Acute Ischemic Stroke. *Brain Sci*. 2020;10(9):641. doi:10.3390/brainsci10090641

13. Hecht N, Schrammel M, Neumann K, et al. Perfusion-Dependent Cerebral Autoregulation Impairment in Hemispheric Stroke. *Ann Neurol*. 2021;89(2):358-368. doi:10.1002/ana.25963
